# Combinatorial delivery of low-dose irradiation and immunotherapy to patients with immune-excluded tumors effectively enhances CD8^+^ T cell functionality

**DOI:** 10.1101/2025.05.12.25327418

**Authors:** Maria Ochoa-de-Olza, Nicolas Rayroux, Martina Imbimbo, Angela Orcurto, Noémie Fahr, Fabrizio Benedetti, Julien Dagher, Giulia Spagniol, David Barras, Eleonora Ghisoni, Blanca Navarro, Arthur Mulvey, Dominik Berthold, Apostolos Sarivalasis, Khalil Zaman, Athina Stravodimou, Antonia Digklia, Rafael Duran, Clarisse Dromain, John O. Prior, Niklaus Schaeffer, Stefan Zimmermann, Michel Obeid, Nikos Chalkidis, Tsourti Zoi, Bettina Bisig, Lionel Trueb, Doga C. Gulhan, Christine Sempoux, Urania Dafni, Stephanie Tissot, George Coukos, Fernanda G. Herrera, Denarda Dangaj Laniti

**Affiliations:** Department of Oncology, Lausanne University Hospital; Ludwig Institute for Cancer Research, Lausanne Branch, University of Lausanne (UNIL); Agora Cancer Research Center, Lausanne, Switzerland; Immuno-oncology Service, Department of Oncology, Lausanne University Hospital, Switzerland; Medical Oncology Service, Department of Oncology, Lausanne University Hospital, Lausanne, Switzerland; Institute of Pathology, Unit of Translational Onco-pathology, Lausanne University Hospital, Lausanne, Switzerland; Department of Gynecology and Obstetrics, Division of Women and Children, Padova University Hospital, Padova, Italy; Department of Radiology and Interventional Radiology, Lausanne University Hospital, Lausanne, Switzerland; Department of Nuclear Medicine and Molecular Imaging, Lausanne University Hospital and University of Lausanne, Lausanne, Switzerland; Service of Immunology and Allergy, Lausanne Center for Immuno-Oncology Toxicities LCIT, Lausanne University Hospital, Lausanne, Switzerland; Scientific Research Consulting Hellas, Statistics Center, Athens, Greece; Institute of Pathology, Lausanne University Hospital and Lausanne University, Lausanne, Switzerland; Krantz Center for Cancer Research, Massachusetts General Hospital, Boston, USA Department of Medicine, Harvard Medical School, Department of Medicine, USA Broad Institute of Massachusetts Institute of Technology and Harvard University, Cambridge, MA, USA; Faculty of Nursing, National and Kapodistrian University of Athens, Athens, Greece. Radiation Oncology; Radiation Oncology Service, Department of Oncology, Lausanne University Hospital, Lausanne, Switzerland

**Keywords:** low dose radiation, PD1, CTLA4, T cells, tumor microenvironment, Homologous recombination repair deficiency

## Abstract

Immune-checkpoint blockade (ICB) has shown significant efficacy across various tumor types. However, tumors with low intraepithelial T-cell infiltration, often referred to as “cold” tumors, are expected to yield poor responsiveness to ICB. We investigated the potential of low-dose radiotherapy (LDRT) to enhance ICB responses in 25 patients with multimetastatic immune-excluded solid tumors through a multi-cohort phase I clinical trial (RACIN). Primary endpoint was to determine the safety and tolerability of the combination of a backbone treatment, comprising nivolumab, ipilimumab, aspirin/celecoxib, and low-dose cyclophosphamide (Cy) in association with escalated LDRT. Secondary endpoints included among others disease control rate (DCR) and overall survival (OS). Exploratory endpoints included biomarkers and molecular correlates of response. The combination treatment showed a manageable safety profile, with Grade 3 or higher adverse events in 12% to 21% of patients across cohorts. The overall DCR was 42%. Progression free survival (PFS) across all cohorts was 2.1 months (95% C.I.: 1.8 – 4.2 months), with the highest PFS observed in cohort 1 which received 0.5 Gy (5.7 months, (95% C.I.: 1.9 - 11.3 months). Median OS was 14.0 months (95% CI: 8.5–24.6 months), with one patient with ovarian cancer still maintaining a complete response at three years follow-up. Site-paired tumor biopsies collected for each patient at baseline and after LDRT +/− Cy enabled the comprehensive characterization of the dynamics of excluded tumor microenvironments (TME) at the single cell level. Response to LDRT and ICB was associated with DNA damage and repair responsiveness and the presence of detectable intratumoral PD1^+^CD8^+^ tumor infiltrating lymphocytes (TILs) at baseline. Our data revealed that LDRT amplified CD8^+^ TIL functionality in responding patients offering mechanistic insights on how LDRT improves ICB effectiveness. In contrast, we observed a radiosensitivity of TILs in tumors of non-responders. Detailed single cell immune profiling before LDRT also highlighted a lack of key immune stimulatory myeloid cells that can therefore limit ICB efficacy in excluded tumors. Collectively, this study represents the most comprehensive profiling of longitudinal samples of cancer patients treated with LDRT. Our findings highlight several genetic, transcriptomic and TME parameters associated with response to combinatorial LDRT and ICB in advanced immune-excluded solid cancers, generating rationale for their validation in larger cohorts.

**One sentence summary:** Combinatorial treatment with low-dose radiotherapy and immune checkpoint blockade in patients with immune-excluded tumors enhances CD8⁺ T cell functionality, particularly in those with DNA repair deficiencies, offering new biomarkers for patient selection.

## INTRODUCTION

Immunotherapy with immune-checkpoint blockade (ICB) targeting CTLA-4, PD1/(L)-1 and LAG-3, has shown clinical efficacy across different tumor types (1, 2). However, many patients experience disease progression, even after initial responses. Tumor response to ICB depends on the composition of the tumor microenvironment (TME) and its immune contexture (3). Expectedly, patients with low T-cell infiltration, so-called “cold” tumors, seldom respond to ICBs (4). As a result, substantial research has focused on strategies to induce inflammation within the TME to enhance tumor responsiveness to ICB (5).

Radiation therapy (RT), particularly at high doses, has emerged as a promising immunomodulatory strategy. High-dose RT (HDRT) acts as an *in situ* vaccine by exposing tumor-specific antigens (6), inducing damage-associated molecular patterns (7), upregulating major histocompatibility (MHC) class I molecules (8), and activating DNA-sensing pathways that stimulate type-I interferon (IFN) production (9). These mechanisms collectively enhance innate and adaptive immune responses, forming the foundation for the abscopal effect - tumor regression at sites distant from the irradiated region (10). However, consistent clinical success in leveraging abscopal effects remains limited, highlighting the need to optimize dose, delivery, sequencing, and combinatorial strategies (11).

Emerging evidence underscores the role of RT parameters in shaping immune responses. Fractionated regimens, such as 8 Gy × 3, mitigate suppressive feedback mechanisms, like TREX1-mediated inhibition of the cyclic GMP-AMP synthase-stimulator of interferon genes (cGAS-STING)-IFN-β pathway, thereby enhancing systemic immunity (12). Conversely, irradiation of tumor-draining lymph nodes can impair T-cell priming, emphasizing the critical importance of precise immune-driven RT delivery (13). Timing is equally critical, with immunotherapies such as anti-PD1 (14) and OX-40 agonists (15) showing greater efficacy when administered after HDRT.

Low-dose radiotherapy (LDRT) at doses below 2 Gy offers an alternative for TME reprogramming. Unlike HDRT, LDRT does not directly cause tumor cell death (16). Instead, LDRT triggers the release of pro-inflammatory cytokines, altering endothelial adhesion receptor expression, and enhancing immune cell trafficking and activation (17). In a preclinical RT5 model of neuroendocrine pancreatic cancer, LDRT induced macrophage polarization toward a pro-inflammatory M1 phenotype and facilitated T cell recruitment through vasculature remodeling (18). Combining HDRT to primary tumors with LDRT to metastatic sites in a lung cancer mouse model further downregulated tumor growth factor (TGF)-β, reprogrammed macrophages, and amplified innate and adaptive immunity, leading to significant tumor regression (19).

Recent studies employing brachytherapy to deliver HDRT and LDRT within the same tumor have demonstrated that dose heterogeneity enhances abscopal effects. High-dose regions promoted MHC class I expression, while low- and moderate-dose regions preferentially supported CD8^+^ T cell recruitment. Interestingly, regulatory T cells (CD4^+^FOXP3^+^) were enriched in high-dose areas. By contrast, uniform RT (2, 8, or 20 Gy) failed to significantly alter immune cell infiltration.

More recently, retrospective clinical data and preclinical models have suggested that intestinal low-dose radiotherapy (iLDRT) enhances the response to immune checkpoint blockade via modulation of gut microbiota (20). This effect is currently being evaluated in a Phase 2 prospective trial testing iLDRT in combination with anti-PD-L1 but without cancer-targeted radiotherapy, in patients progressing on immunotherapy (20). Preliminary clinical data suggest that iLDRT may enhance systemic immune responses and improve tumor control rates in select patients, warranting further investigation in larger cohorts.

Our group explored the potential of LDRT in reprogramming the TME and enhancing the immunotherapy response in a syngeneic ID8 mouse model of metastatic ovarian cancer, characterized by low T cell infiltration and resistance to immunotherapy. Low-dose whole-abdominal irradiation (0.5–2 Gy) increased lymphocytes, monocytes, and NK cells in tumors, along with a favorable CD8^+^ T cell to regulatory T cell (Treg) ratio. Genomic analysis revealed upregulation of immune checkpoint genes (PD-1, CTLA-4), the Treg marker FOXP3, and the myeloid receptor CD40. We developed a combinatorial radio-immunotherapy strategy (RACIM, **Ra**diotherapy **C**ombinatorial **Im**munotherapy) with LDRT (1 Gy), low-dose cyclophosphamide, anti-PD1, anti-CTLA-4, and a CD40 agonist. RACIM achieved an 83.5% tumor response and a 15% cure rate in mice bearing ID8 ovarian cancers with all treatment components essential for survival benefits. RACIM reprogrammed macrophages to an anti-tumoral M1 phenotype and induced dendritic cell activation and priming of tumor-specific CD4^+^ and CD8^+^ T cells expressing IFN-γ, granzyme B (GrB), and perforin, through RAE-NKG2D crosstalk (16).

Based on these findings, we designed a dose-escalation Phase I first-in-human clinical trial (NCT03728179) involving patients with cold, non-immune-infiltrated solid tumors. Patients received LDRT (0.5-1 Gy) to large fields of metastatic disease, paired with an immunotherapy approach similar to the orthogonal strategy used in the mouse model. Pre- and post-LDRT tumor biopsies were collected from the same irradiated area to assess the steady-state immune context and explore the dynamics of radiation-induced immune reprogramming. This approach aimed to uncover key mechanisms driving T cell–mediated tumor rejection and therapeutic resistance, as well as identify molecular markers to guide future therapeutic strategies.

## RESULTS

### Patient characteristics

Between March 2019 and September 2021, 25 patients with metastatic TIL-negative solid tumors, defined by a median of fewer than 5 intraepithelial TILs per high power field (HPF) as determined by a clinical pathologist, were enrolled in the multi-cohort, open label prospective phase I clinical trial combining low dose **<u>r</u>**adiotherapy, **a**spirin/celecoxib, low dose **<u>c</u>**yclophosphamide (Cy), **i**pilimumab and **<u>n</u>**ivolumab (RACIN) (**Fig. 1A, Supp. 1A-C**, **Methods**). The cohort distribution included three patients in cohort 1, six in cohort 2, six (randomized) in cohort 3A, and ten (randomized) in cohort 3B. Baseline demographics and disease characteristics are summarized in Table 1. Overall, 52% of patients were male, with median age of 66 years (range: 19-82), and 72% had Eastern Cooperative Oncology Group (ECOG) performance status 0. Tumor mutational burden (TMB) ranged from 0 to 7.8 mutations per megabase (Mb) (average: 3.0 mutations/Mb), and PD-L1 tumor proportion score (TPS) was < 1% in 64% of the patients.

**Fig. 1:**
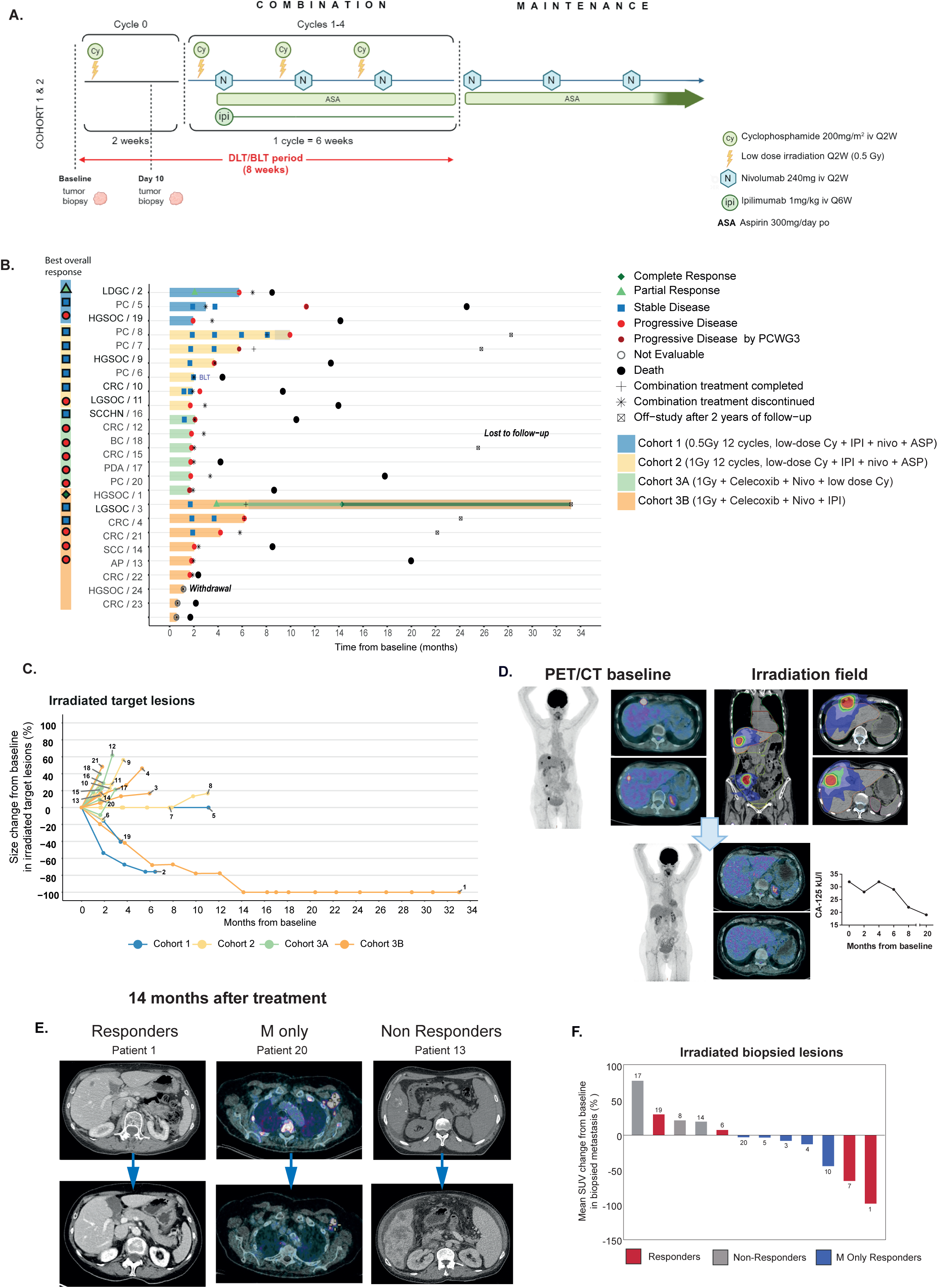
Study outcomes: Safety and Efficacy. **(A)** Therapeutic schema of the phase I clinical trial RACIN for cohort 1 and 2. **(B)** Swimmer plot shows treatment duration, type of response (in accordance with RECIST 1.1 and PCWG3 for prostate cancer) and response duration. Bar length indicates time to treatment failure. The shaded part of the bar indicates time on nivolumab maintenance. **(C)** Spiderplot depicts size change from baseline in irradiated target lesions (as per RECIST criteria) measured by CT scan, following treatment with LDRT +/− Cy and ICB**. (D)** Left panel: baseline PET/CT of a HGSOC patient, with liver metastases and pelvic implants. Right panel: irradiation fields covering all visible metastasis. Bottom panel: CR after 14 months of treatment, with disappearance of liver metastases and pelvic implants. Line chart displaying tumor marker CA-125 levels since the start of treatment in the HGSOC patient with CR. **(E)** Representative CT scan images (left and right) and PET/CT scan images (middle) of each response category**. (F)** Waterfall plot shows mean SUV change from baseline in biopsied metastases as measured by PET/CT, following treatment with LDRT +/− Cy and ICB. LDGC: low differentiated gallbladder carcinoma. HGSOC: high grade serous ovarian carcinoma. PC: prostate adenocarcinoma. CRC: Colorectal adenocarcinoma. LGSOC: low grade serous ovarian carcinoma. SCC: Squamous cervical carcinoma. SCCHN: squamous cell carcinoma of the head and neck. PDA: pancreatic ductal adenocarcinoma. BC: ductal breast carcinoma. AP: appendix adenocarcinoma.

**Table 1:** Patient and tumor baseline characteristics, by arm/cohort and overall.

|  | Cohort 1<br>(n= 3) | Cohort 2<br>(n= 6) | Cohort 3A<br>(n= 6) | Cohort 3B<br>(n= 10) | Overall<br>(N= 25) |
| --- | --- | --- | --- | --- | --- |
| **Gender - n (%)** |  |  |  |  |  |
| Female | 1 (33.3) | 2 (33.3) | 4 (66.7) | 5 (50) | 12 (48) |
| Male | 2 (66.7) | 4 (66.7) | 2 (33.3) | 5 (50) | 13 (52) |
| **Age** |  |  |  |  |  |
| Median (range) | 59, 70, 70 | 66 (19 - 72) | 56.5 (32 - 82) | 63 (40 - 78) | 66 (19 - 82) |
| **ECOG - n (%)** |  |  |  |  |  |
| 0 | 2 (66.7) | 3 (50) | 6 (100) | 7 (70) | 18 (72) |
| 1 | 1 (33.3) | 3 (50) | - | 3 (30) | 7 (28) |
| **Type of cancer - n (%)** |  |  |  |  |  |
| Prostate adenocarcinoma | 1 (33.3) | 3 (50) | 1 (16.7) | 1 (10) | 6 (24) |
| High grade serous ovarian carcinoma | 1 (33.3) | 1 (16.7) | - | 2 (20) | 4 (16) |
| Low grade serous ovarian carcinoma | - | 1 (16.7) | - | 1 (10) | 2 (8) |
| Adenocarcinoma of the appendix | - | - | - | 1 (10) | 1 (4) |
| Colorectal adenocarcinoma | - | 1 (16.7) | 2 (33.3) | 4 (40) | 7 (28) |
| Squamous cell carcinoma of the head and neck | - | - | 1 (16.7) | - | 1 (4) |
| Low differentiated gallbladder carcinoma | 1 (33.3) | - | - | - | 1 (4) |
| Ductal breast carcinoma | - | - | 1 (16.7) | - | 1 (4) |
| Pancreatic ductal adenocarcinoma | - | - | 1 (16.7) | - | 1 (4) |
| Squamous cell cervical carcinoma | - | - | - | 1 (10) | 1 (4) |
| **PD-L1 TPS, n (%)\*** |  |  |  |  |  |
| < 1% | 2 (6) | 6 (100) | 3 (50) | 5 (50) | 16 (64) |
| 1-49% | 1 (33.3) |  | 1 (16.6) |  | 2 (8) |
| ≥ 50% |  |  |  |  |  |
| Unknown |  |  | 2 (33.3) | 5 (50) | 7 (28) |
| **Stage at study entry - n (%)** |  |  |  |  |  |
| IV | 3 (100) | 4 (66.7) | 4 (66.7) | 2 (20) | 13 (52) |
| IVB | - | 2 (33.3) | 1 (16.7) | 5 (50) | 8 (32) |
| IVC | - | - | 1 (16.7) | 3 (30) | 4 (16) |
| **No. of prior lines of treatment - n (%)** |  |  |  |  |  |
| 2 lines | 1 (33.3) | - | 3 (50) | 2 (20) | 6 (24) |
| ≥ 3 lines | 2 (66.6) | 6 (100) | 3 (50) | 8 (80) | 19 (76) |
| **Prior radiotherapy - n (%)** |  |  |  |  |  |
| Yes | - | 3 (50) | 5 (83.3) | 6 (60) | 14 (56) |
| No | 3 (100) | 3 (50) | 1 (16.7) | 4 (40) | 11 (44) |

\* PD-L1 status was determined by immunohistochemistry using the Ventana PD-L1 (SP263) clone.

The patients included had a wide range of metastatic solid tumors and had received a median of three previous lines of treatment prior to enrollment, ranging from 2 to 7 lines of treatment, with 52% of patients having received four or more lines of treatment. Fifty-six percent of the patients had undergone prior RT and re-irradiation of the same previously irradiated lesions was not allowed.

### Primary outome: Safety

Cohort 1 followed a 3+3 algorithm adapted for multidrug combination therapy, with all three patients enrolled either simultaneously or sequentially, without any waiting period between enrollments (**Supp. 1D)**. As no dose limiting toxicities (DLTs) or bone limiting toxicities (BLTs) were observed in the three enrolled patients in Cohort 1, an additional six patients were recruited into Cohort 2. There were 17% ≥ grade 3 (G3) adverse events (AEs) in cohort 1, and 13% in cohort 2. Immune-related serious AEs of G3 or more (not constituting DLT or BLT) occurred in three patients (33%) in cohort 1 and 2. A BLT was recorded in cohort 2. Most frequent treatment related toxicities in the DLT/BLT period in were lymphocyte count decrease (46%), fatigue (50%) and nausea (46%) (**Supp. 1E**).

In Cohort 1, a prostate cancer (PCa) patient developed recurrent, refractory G3 colitis requiring high-dose steroids, infliximab and vedolizumab for management. This AE was attributed to the combination of ipilimumab and nivolumab, treatment discontinuation was necessary, and the disease subsequently progressed. In Cohort 2, one PCa patient experienced G4 myocarditis, necessitating tocilizumab treatment and resulting in treatment discontinuation and progressive disease. Another patient in cohort 2 with PCa developed G3 immune-related hepatitis which responded rapidly to corticosteroids; patient was able to continue treatment without ipilimumab, obtaining metabolic response as per ^68^Ga-PSMA-PET, ultimately progressing after seven months of treatment.

Both G3 colitis and G4 myocarditis proved refractory to corticosteroid therapy, significantly impacting the patients’ lives. Consequently, the investigators attributed the recalcitrant and persistent nature of the toxicity to the concomitant administration of Cy and ipilimumab, as previous studies (21, 22) demonstrated that this combination significantly decreases systemic immune suppressive T regs. As a result, and despite not meeting the formal criteria for trial termination, the decision was made, in consultation with the Independent Data Safety Monitoring Committee, to amend the study protocol. Patients in subsequent Cohort 3 were thus randomized to receive either Cy or ipilimumab, in addition to the rest of the combination therapy (**Supp. 1A, B**). No DLTs related to LDRT were reported in cohort 1 and 2, and therefore the maximum tolerated dose (MTD) of LDRT was not reached, being 1 Gy the recommended dose to be implemented in the randomized cohort 3. Despite observing 12% and 21% of ≥ G3 AEs in Cohorts 3A and 3B, respectively, no DLTs or BLTs were identified in these cohorts.

### Treatment administration

Compliance to treatment is depicted in Supp. Fig 1C. In cohort 1 and 2, all patients completed the DLT/BLT period except for the patient meeting BLT criteria. In cohort 3, 60% of patients did not complete the DLT/BLT period due to PD (7 patients, 47%), investigator decision (1 patient, 7%), one patient withdrew consent (7%) and one patient progressed (7%) before starting treatment. Four patients were replaced in cohort 3B (**Supp. Table 1**).

### Secondary Outcomes: Efficacy

Twenty-four (96%) patients that started treatment were evaluable for efficacy. Clinical response to the combination therapy was assessed by computed tomography (CT) scan every 9 weeks starting C0D1 using Response Evaluation Criteria in Solid Tumors (RECIST 1.1). For PCa patients, response was evaluated according to the Prostate Cancer Working Group 3 (PCWG3) criteria. When available, responses were also assessed using relevant blood tumor markers.

Complete response (CR) and partial response (PR) were observed in 2 out of 24 patients: one patient with high grade serous ovarian cancer (HGSOC, patient 1) in cohort 3B and one patient with gallbladder cancer (GC, patient 2) in cohort 1, respectively (**Table 2**). Eight patients exhibited stable disease (SD) as their best response. These included four with PCa, two with colorectal cancer (CRC), one with low-grade serous ovarian cancer (LGSOC), and one with HGSOC. The duration of disease stabilization for these patients ranged from 11 to 49 weeks. The remaining eleven patients experienced progressive disease. Additionally, three patients did not complete the prescheduled 9-week CT scan due to rapid early progression of the disease, which led to their withdrawal from the study.

**Table 2:** Best overall response (BOR) and objective response rate (ORR) by RECIST 1.1(and by PCWG3 for prostate cancer), by arm/cohort in the efficacy population (n = 24)

|  | **Cohort 1**<br>**(n= 3)** | **Cohort 2**<br>**(n= 6)** | **Cohort 3A**<br>**(n= 6)** | **Cohort 3B**<br>**(n= 9)** |
| --- | --- | --- | --- | --- |
| **Best overall response (BOR)** |  |  |  |  |
| Complete Response (CR) | - | - | - | 1 (11.1%) |
| Partial Response (PR) | 1 (33.3%) | - | - | - |
| Stable Disease (SD) | 1 (33.3%) | 5 (83.3%) | - | 2 (22.2%) |
| Progressive Disease (PD) | 1 (33.3%) | 1 (16.7%) | 6 (100.0%) | 3 (33.3%) |
| Not evaluable (NE) | - | - | - | 3 (33.3%)<sup>a</sup> |
| **Objective Response<sup>b</sup> Rate (ORR)** |  |  |  |  |
| ORR (95% binomial C.I.) | 33.3% (0.8% - 90.6%) | 0.0% (0.0% - 45.9%)<sup>c</sup> | 0.0% (0.0% - 45.9%)<sup>c</sup> | 11.1% (0.3% - 48.2%) |

(a) All 3 patients discontinued early in the course of trial and were replaced. No follow-up tumour assessments available.
(b) Complete or partial response.
(c) 95% Clopper Pearson exact C.I.

The overall response rate (CR + PR) was 8%, and DCR was 42% (**Table 2, Supp. Table 2**). A comprehensive evaluation of all patients is illustrated in Fig. 1B. Seventy-eight percent of target and non-target lesions were irradiated (range: 16-100%), and the change in percentage of all irradiated target lesions compared to baseline throughout the study is depicted in Fig. 1D. Noteworthy cases with significant outcomes are described in more detail below.

The patient with HGSOC who achieved a CR (patient 1) was treated in cohort 3B. She received LDRT 1 Gy, celecoxib, ipilimumab, and nivolumab, omitting Cy as per protocol. She completed the study and received 1 year of maintenance nivolumab, which she continues to receive through compassionate use. This patient achieved 20% tumor shrinkage of target lesions according to RECIST v.1.1 at the first scheduled CT scan, accompanied with a 19% reduction in CA-125 tumor marker after 9 weeks on treatment (**Fig. 1C**). She finally achieved a CR 14 months after the start of treatment, and she continues to tolerate nivolumab maintenance well and is now over one year outside of the study, continuing nivolumab off-trial.

The patient with LDGC who achieved a PR in cohort 1 entered the study after two lines of treatment and received LDRT 0.5 Gy, aspirin, ipilimumab, nivolumab and Cy. He showed a decrease in both irradiated and non-irradiated metastases at first CT scan after 9 weeks of treatment initiation, but ultimately progressed due to the development of new lesions outside the irradiated area.

Interestingly, a PCa patient obtaining SD as per PCWG3 criteria demonstrated a dramatic metabolic response in fields that received LDRT based on molecular imaging with Gallium-68 prostate-specific membrane antigen positron emission tomography (^68^Ga-PSMA-PET). The patient ultimately presented PD related to new lesions emerging uniquely outside of the irradiated areas.

At data cut off, 11^th^ of December 2023, median time to treatment failure (TTF) was 1.9 months (95% C.I.: 1.7 – 3.0) (**Supp. 1F**). Median progression free survival (PFS) for all cohorts was 2.1 months (95% C.I.: 1.8 – 4.2 months) (**Supp. 1G**), with the highest PFS observed in cohort 1 (5.7 months, (95% C.I.: 1.9 - 11.3 months), and the lowest observed in cohort 3A (**Supp. Table 3**) with a median overall survival (OS) for all cohorts of 14.0 months (95% C.I.: 8.5 month – 24.6) (**Supp. 1H, Supp. Table 4**).

### Exploratory outcomes

#### Biopsied metastases responses

Each patient had two biopsies performed on a single irradiated metastasis: one prior to irradiation (pre-treatment) and a second biopsy 10 days following LDRT (+/− Cy), but before the initiation of ICB (**Supp. Fig 2A** and **Supp. Fig 2B**). The same irradiated metastasis was biopsied at both timepoints in all patients, except for patient 3 which was biopsied in a different irradiated metastases for technical reasons. To assess the biological effects of LDRT and ICB *in situ*, we quantified the reduction in size of each biopsied and irradiated metastasis obtained by CT scans, and employed them to stratify responses, instead of using RECIST responses. This approach was chosen given that only two objective responses were observed, which posed limitations in performing translational research. Overall, the biopsied metastases reflected the general disease evolution observed in irradiated and non-irradiated metastases with the exception of one patient (patient 17) who exhibited a slight reduction in the size of some metastases, though not in the metastasis that was biopsied. Consequently, this patient was not classified as a “Responder”.

We observed that 5 patients (21%) presented a reduction in the size of their biopsied and irradiated metastasis (**Fig. 1D**), and we defined this as an *in situ* responding category, classifying this group of patients as “Responders” (R) (**Fig. 1D, E**). In addition, we assessed metabolic changes by PET/CT in biopsied and irradiated metastasis and observed a decrease in mean standard uptake value (SUV) following treatment in 7 out of the 12 patients (58%) for whom PET/CT was available (**Fig. 1F**). Moreover, we observed that 5 out of these 7 patients exhibited a reduction in mean SUV but without a reduction in the size of the metastasis by CT scan. This pattern was classified as a distinct response category termed metabolic “M-only” Responders (21%) (**Fig. 1E, F**). Patients with biopsied and irradiated metastasis not meeting the criteria of “Responders” or “M-Only” Responders were categorized as “Non-Responders” (NR, 11 patients, 46%) (**Fig. 1E**).

### TME analysis and association between TME and efficacy

#### DNA repair pathway signatures can predict response to LDRT and ICB

As previously described, patients were enrolled based on the absence of intraepithelial T cells. Consistent with this criterion, subsequent analysis revealed that these tumors exhibited an immune-excluded phenotype, with a significantly higher density of CD8⁺ T cells localized in the stromal compartment compared to the intraepithelial region (**Supp. Fig 2C**). This pattern—akin to what has been variably described as a ‘cold’ or ‘immune desert’ phenotype—is infrequently observed in clinical settings, reflecting the evolving understanding of tumor immune landscapes (23).

To further understand the characteristics of immune-excluded tumors and their TME in relation to their response to LDRT, we conducted a comprehensive TME analysis that included immunohistochemistry (IHC), multiplex immunofluorescence (mIF), bulk ribonucleic acid (RNA), and single-cell RNA sequencing (**Supp. Fig 2A**).

Principal Component Analysis (PCA) of bulk RNAseq from the complete cohort identified tumor type and biopsy location site as significant variables influencing the analysis. Consequently, and considering sample availability, we focused our analysis on ovarian and PCa samples representing the majority of tumor histologies (further details provided in Methods section). Reactome pathway analysis on baseline HGSOC and PCa samples revealed elevated signature scores for DNA damage response and repair pathways in lesions that regressed following LDRT. This was observed in both radiological “responders” (Rs) and those classified as “metabolic only” responders (**Fig. 2A-D**, **Supp**. **2D**, **E**). Interestingly, the G2M checkpoint pathway was significantly upregulated in Rs but not in M-only or NRs lesions (**Fig. 2E**). In contrast, pathways suggestive of epithelial-mesenchymal transition (EMT) were not different among response categories, albeit a higher trend being observed in NRs (**Supp**. **2F**).

**Fig. 2:**
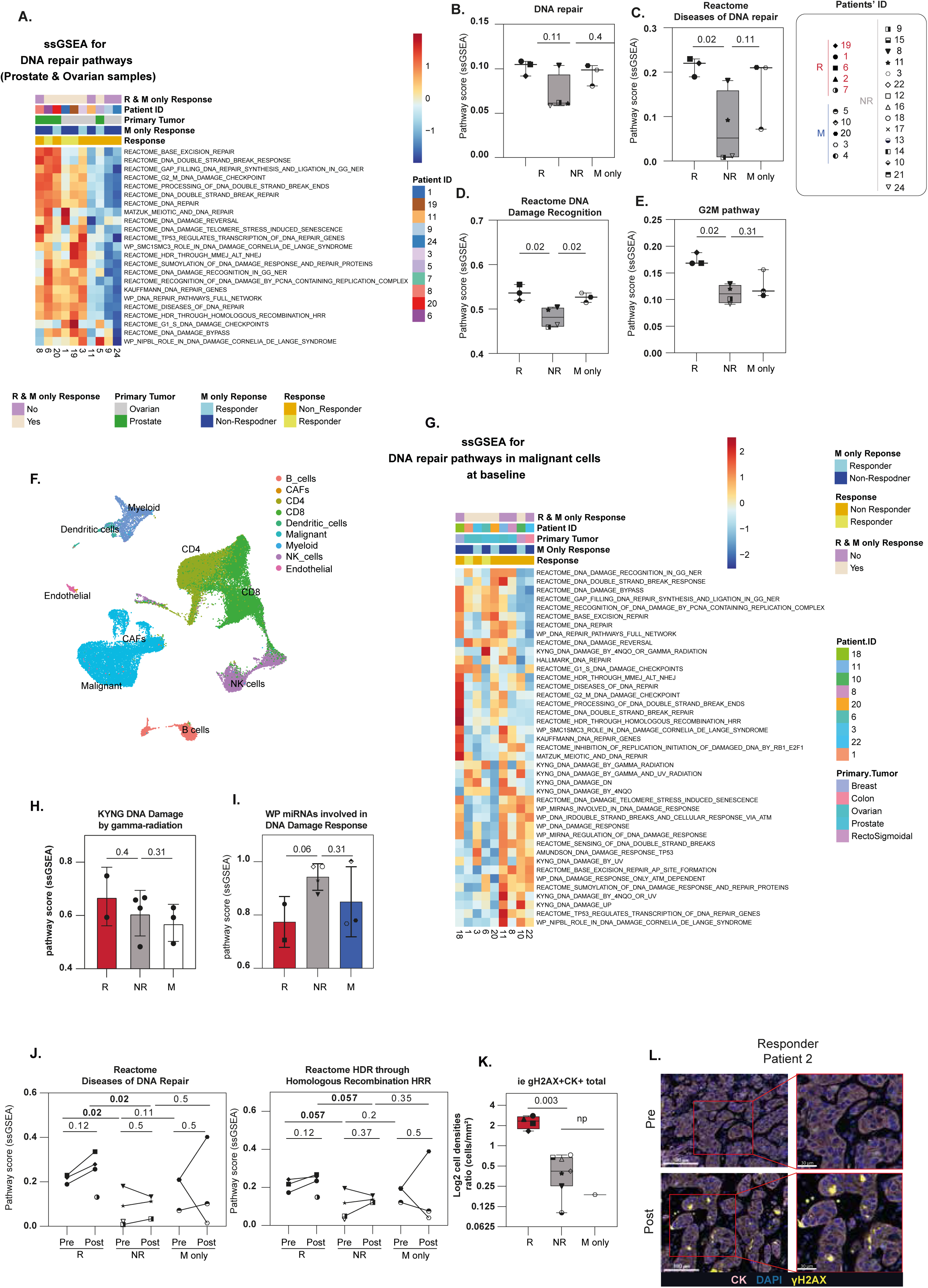
DNA repair pathway signatures can predict response to Low Dose Irradiation and Immune checkpoint blockade. **(A)** Heatmap showing select DNA repair pathways in ovarian and prostate samples at baseline (see Methods). The heatmap displays ssGSEA scores for reactome DNA repair pathways from bulk RNA sequencing. Box plots displaying ssGSEA pathway scores from bulk RNA Seq at baseline for DNA repair pathways **(B-D)** and G2M pathway **(E)**. **(F)** UMAP projection showing clustering of major cell populations from viable-cell sorted cells of ten baseline and eight post-treatment tumors, with seven of them being paired. **(G)** Heatmap showing ssGSEA scores for DNA repair pathways in malignant cells from scRNA sequencing available at baseline. Box plots displaying ssGSEA pathway scores from scRNA Seq at baseline for DNA Damage by gamma-radiation **(H)** and miRNAs involved in DNA Damage Response **(I)** (Mann-Whitney, one-tailed). **(J)** Line graphs showing ssGSEA pathway scores from bulk RNA Seq at baseline and at day 10 for different DNA repair signatures (paired: Wilcoxon test; unpaired: Mann-Whitney test; one tailed). **(K)** Boxplot displaying the log2 ratio of γH2AX^+^CK^+^ cell densities of cells post-treatment compared to pre-treatment (0YL1 (M only responder) is not displayed as there was fluorescence overlap between γH2AX and CD11c) (Mann-Whitney, one-tailed). **(L)** Multiplex immunofluorescent images displaying γH2AX^+^CK^+^ cells at baseline and 10 days after LDRT delivery in the patient with LDGC and partial response.

We performed scRNAseq analysis on a subset of patient biopsies where tissue quality and quantity were sufficient and documented the tumor and TME atlas of immune-excluded solid tumors (**Fig. 2F**). ScRNAseq analysis of the malignant compartment at baseline corroborated these findings (**Fig. 2G, H**), further revealing that NRs upregulated the expression of regulatory miRNAs signatures involved in DNA damage and potentially its suppression (**Fig. 2I**). Moreover, Rs displayed increased DNA repair responses at baseline, which were further augmented following LDRT treatment. In contrast, a heterogeneous response was observed among M-only responders (**Fig. 2J, Supp. 2G**). This finding was corroborated at the protein level, as biopsies from Rs showed an increased fold change in the DNA damage marker γH2AX after LDRT treatment. (**Fig. 2K, L**).

This was corroborated by DNA sequencing (based on a targeted high-throughput sequencing (HTS) panel covering 400 genes) which revealed an association between mutations in DNA damage repair genes, such as *ATM*, in 2 out of 4 patients who exhibited an increase in γH2AX after LDRT +/− Cy (**Supp. 3**). Additionally, we observed a higher frequency of mutations in genes related to chromatin modification and transcription regulation, such as *KMT2A* and *TRRAP*, associated with a γH2AX increase *in situ*. These mutations could potentially disrupt gene expression required for DNA repair and cell cycle control, exacerbating the accumulation of DNA damage and sensitizing tissues to LDRT (**Supp. 3**)

Additionally, the HGSOC patient who achieved CR (**Fig. 1C**) presented with mutations in *CDK12*, *TP53* and *ATM*. This patient also exhibited a high level of large-scale state transitions (LST), a marker of genomic instability and had a TMB of 7.8 mutations/Mb as per the 400-genes HTS panel. Further targeted DNA sequencing applying the SigMA algorithm (24) revealed the presence of mutational signature 3, which is associated with homologous recombination deficiency (HRD) (25).

#### Immune-cell atlas of immune-excluded tumors and their role for response to LDRT and ICB

Single cell analysis of the immune TME using mIF and scRNAseq revealed that in excluded tumors of RACIN patients, even the mere presence of 1 or 2 intraepithelial PD1^+^CD8^+^ per mm^2^, presumably tumor specific T cells (26), was significantly associated with favorable tumor responses to LDRT and ICB (**Fig. 3A, B, Supp. 4A-C**).

**Fig. 3.**
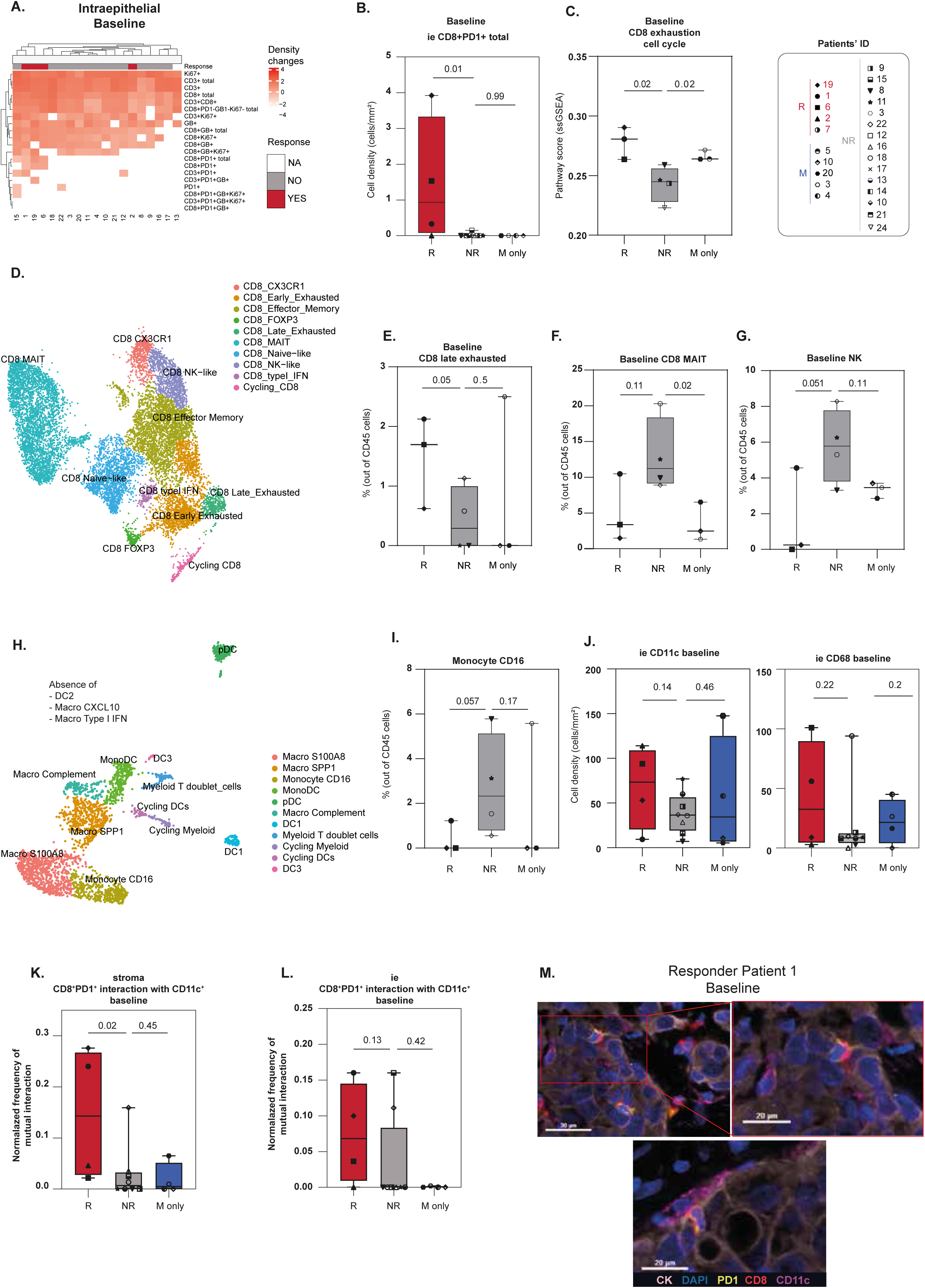
Immune-cell atlas of immune-excluded tumors and their role for response to Low-Dose Irradiation and immune checkpoint blockade. **(A)** Heatmap of intraepithelialcell densities quantified by mIF of different T cell phenotypes in baseline tumor biopsies. **(B)** Box plot displaying cell density (cells/mm^2^) of intraepithelial CD8^+^PD1^+^ total cells at baseline by mIF. (Mann-Whitney, two-tailed). **(C)** Box-plot showing pathway score (ssGSEA) from bulk RNAseq of the pathway CD8^+^ exhaustion and cell cycle at baseline (Mann-Whitney, one-tailed). **(D)** UMAP projection displaying sub-clustering of CD8^+^ T cells from viable-sorted data of ten baseline and eight post-treatment tumors. Box-plot displaying proportion of CD8^+^ late exhausted **(E)** and MAIT CD8^+^ T cells **(F)** in the CD45^+^ cells at baseline (Mann-Whitney, one-tailed). **(G)** Boxplot displaying proportion of NK cells in the CD45^+^ cells at baseline (Mann-Whitney, one-tailed). **(H)** UMAP projection displaying sub-clustering of myeloid cells from viable-sorted data of ten baseline and eight post-treatment tumors. **(I)** Boxplot displaying proportion of monocyte CD16 cells in the CD45^+^ cells at baseline (Mann-Whitney, one-tailed). **(J)** Box-plots displaying cells densities (cells/mm2) of myeloid cells at baseline by multiplex immunofluorescence (Mann-Whitney, one-tailed) **(K)** Box plots displaying mutual interaction between CD8^+^PD1^+^ T cells with CD11c^+^ cells in the stroma **(K)** and in the tumor **(L)** (Mann-Whitney, one-tailed). **(M)** Multiplex immunofluorescent images displaying CD11c^+^ and CD8^+^ T cell interaction at baseline in the patient with HGSOC achieving CR.

Confirming this observation, R patients exhibited elevated baseline CD8^+^ exhausted and cell cycling signatures at baseline as captured by bulk RNAseq (**Fig. 3C**). We further corroborated these findings by scRNAseq analysis and showed that R patients had higher levels of exhausted CD8^+^ T cells states (**Fig. 3D, E**), consistent with recent descriptions by our group and others (27, 28). The presence of these exhausted CD8^+^ T cells in patients showing local tumor response suggests pre-existing tumor immune recognition through chronic antigen exposure.

Furthermore, we observed that R patients exhibited higher CD8^+^ T cell infiltration in less proliferative tumors (**Supp. 4D**), indicating that lower tumor aggressiveness may permit greater CD8^+^ T cell infiltration in this patient subset. Moreover, R patients showed a higher presence of intraepithelial CD4^+^ T cells at baseline (**Supp. 4E**). Indeed, CD4^+^ T cells can be present within tumors and have demonstrated to contribute to anti-tumor immune responses as also observed in our previous pre-clinical studies (16, 29, 30).

Interestingly we observed that NRs had increased levels of MAIT CD8^+^ T cell states (31) (**Fig. 3F**), NK cell states (**Fig. 3G**) but also CD16^+^ monocytes (**Fig. 3H, I**), suggesting a potential immune non-conducive TME. No significant differences were observed across other CD8^+^ (**Supp. 4F**), CD4^+^ T cell states, (**Supp. 4G, H**), innate T cells (**Supp. 4I, J**), or myeloid cells (**Fig. 3J, Supp. 4K**) across the different response categories. Despite our cohort being immune-excluded and hence not expected to respond to ICB, our findings suggest that even a small number of CD8^+^PD1^+^/exhausted T cells can be sufficient to induce a tumor response (**Supp. 4C**).

To elucidate the cellular crosstalk between myeloid cells and CD8^+^ T cells, we assessed their spatial relationships *in situ* by mIF. We observed a heightened interaction between CD11c^+^ cells and CD8^+^PD1^+^ T cells within the stroma of R patients compared to NRs at baseline (**Fig. 3K**). In contrast, such interactions were less pronounced in tumor islets (**Fig. 3L, and 3M**), suggesting that T cell-myeloid niches present at baseline may play a pivotal role in recruiting and supporting tumor-reactive T cells in responder patients (26, 32).

Notably, our RACIN single-cell atlas revealed a complete absence of co-stimulatory dendritic cell type 2 (DC2), or immunoreactive CXCL9/10^+^ and interferon-stimulated gene positive (ISG^+^) macrophages (**Fig. 3H**), previously identified in inflamed melanomas (28). Indeed, the density of intratumoral DC2 has been shown to correlate with responsiveness to anti-PD-1 therapy (33), while CXCL 9 and CXCL10-secreting macrophages are crucial for effective anti-tumor immune-responses following ICB in melanoma patients (34). The absence of these pro-inflammatory cells suggests that the TME in immune-excluded tumors is highly immunosuppressive, potentially counteracting the efficacy of ICB and precluding LDRT to unleash its full potential.

Overall, our findings provide a comprehensive characterization of T cells and myeloid cells in the TME of immune-excluded tumors, and highlight the absence of pro-inflammatory myeloid cells, offering a novel immune-cell atlas for these tumors.

#### Low dose irradiation increased functionality of tumor-infiltrating CD8^+^ T cells

To assess the impact of LDRT +/− Cy on the TME, we quantified intratumoral immune cell densities using mIF in pre- and post-treatment biopsies. Post-treatment evaluation of the TME focused solely on the effects of LDRT +/− Cy, as ICB was administered after the biopsies. Quantification of intratumoral CD8^+^ cells revealed an increase in the intra-epithelial compartment after LDRT +/− Cy (**Fig. 4A**). Three out of 4 R patients exhibited an increase in intraepithelial CD8^+^ T cells after LDRT +/− Cy (**Fig. 4B**) and R patients demonstrated higher CD8^+^ T cell densities post LDRT +/− Cy treatment compared to NRs (**Fig. 4B**). Independently, patients who showed an increase in intraepithelial CD8^+^ T cells after LDRT +/− Cy were more likely to have mutations in DNA repair and cell cycle related genes (*TP53*, *FANCM*, and *CDK12*, *ATM*) (**Supp. 5A**), suggesting again that tumor cells with impaired DNA repair mechanisms may be more susceptible to LDRT, potentially enhancing immunogenic cell death and subsequently boosting CD8^+^ T cell infiltration and response to ICB.

**Fig 4:**
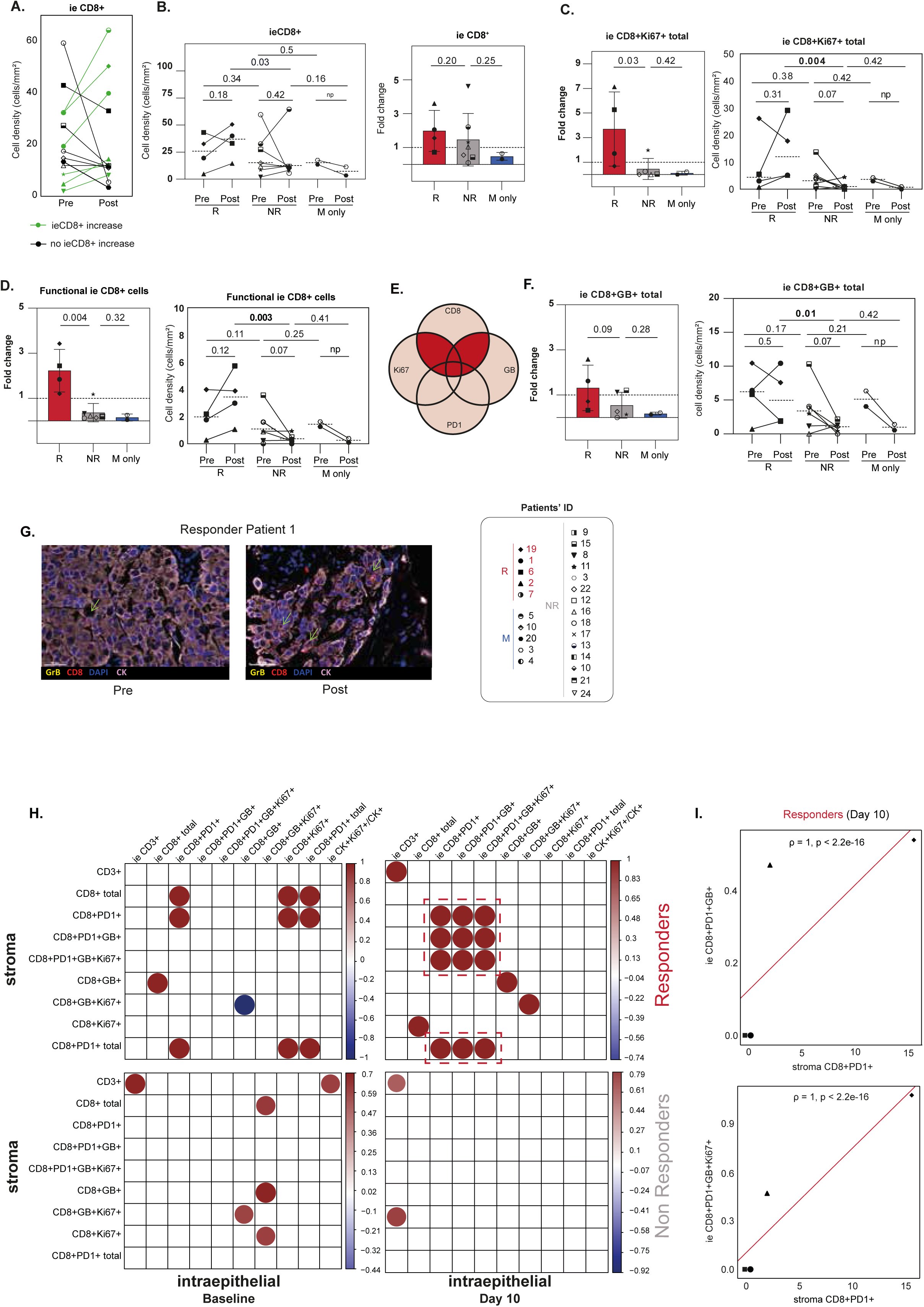
Low dose irradiation increased functionality of tumor-infiltrating CD8^+^ T cells. **(A)** Line graph displaying cell densities (cells/mm2) of intraepithelial CD8^+^ T cells of all patients at baseline and day 10 by mIF**. (B)** Left: Line graph displaying cell densities (cells/mm2) per patient and mean per response category of intraepithelial CD8^+^ cells at baseline and at day 10 by mIF of tissue available at both timepoints (R day 10 versus NR day 10: Mann-Whitney, one-tailed). Right: Box-plot displaying fold change of intraepithelial CD8^+^ cells after LDRT +/− Cy by mIF (Mann-Whitney, one-tailed). **(C)** Left: Box-plot showing fold change of proliferating intraepithelial CD8^+^Ki67^+^ T cells after LDRT +/− Cy by mIF (Mann-Whitney, one tailed). Right: Line graph showing cell densities (cells/mm2) of proliferating intraepithelial CD8^+^Ki67^+^ T cells at baseline and at day 10 by mIF (R day 10 vs NR day 10: Mann-Whitney, one tailed). **(D)** Left: Box plot showing fold change of functional intraepithelialCD8+ T cells after LDRT +/− Cy by mIF (Mann-whitney, one-tailed). Right: Line graph showing cell densities (cells/mm^2^) of functional intraepithelial CD8^+^ T cells at baseline and at day 10 by mIF (R day 10 vs NR day 10: Mann-whitney, one-tailed). **(E)** Venn diagram of CD8^+^ T cell phenotypes. Functional CD8^+^ T cells were defined as those expressing CD8^+^ plus at least one functional marker (dark red). **(F)** Left: Box-plot showing fold change of cytotoxic CD8^+^GB^+^ T cells after LDRT +/− Cy by mIF (Mann-Whitney, one-tailed). Right: Line graph showing cell densities (cells/mm2) of cytotoxic intraepithelial CD8^+^GB^+^ T cells at baseline and at day 10 by mIF (R day 10 vs NR day 10: Mann-Whitney, one tailed). **(G)** Multiplex immunofluorescent images displaying cytotoxic CD8^+^ T cells at baseline and 10 days after LDRT delivery in the patient with HGSOC who achieved CR. **(H)** Correlation map of cell densities by mIF (Spearman’s rank correlation): x-axis: cell phenotypes in the stroma, y-axis: intra-epithelial cell phenotypes. Only significant correlations with p < 0.05 are plotted. Responders are plotted on the upper graphs; non-responders are plotted in the lower graphs. Dashed lines highlight cell phenotype correlations that appear in responder patients on day 10 after treatment, which are neither present baseline nor in non-responder patients at any timepoint. **(I)** Representative correlation graphs (Spearman’s rank correlation) of stromal and intraepithelial CD8^+^ cell phenotypes which appear on day 10 in responder patients.

We next examined LDRT treatment-induced changes in CD8^+^ T cell functionality. Whole-slide mIF analysis revealed that R patients exhibited a significant increase in proliferative intraepithelial Ki67^+^CD8^+^ cells (**Fig. 4C, Supp. 5B**), displaying polyfunctional effector properties, as evidenced by their coexpression of PD1, and/or GrB following LDRT +/− Cy (**Fig. 4D-F Supp. 5C, D**), and as exemplified in the case of our CR HGSOC patient who received LDRT in the absence of Cy (**Fig. 4G**). In contrast, NR patients showed a trend toward decreased polyfunctionality and proliferation of CD8^+^ T cells following LDRT +/− Cy (**Fig 4. C, D, F**), a phenomenon not observed in R patients.

These findings suggest that LDRT can reinvigorate CD8^+^ exhausted T cells in patients with immune-excluded tumors who subsequently responded to ICB. Our results also indicate distinct CD8^+^ TIL states between R and NR (**Fig 3D-F**), which may also highlight differences in radiosensitivity. For example, naïve T cells could be more radio-sensitive than memory T cells (35). However, our scRNA seq did not reveal significant differences in apoptosis pathways between activated and non-activated CD8^+^ TILs after LDRT +/− Cy (**Supp. 5E**), although this could be potentially limited by the low number of scRNA samples and the low number of CD8^+^ T cells detected.

Further analysis demonstrated that R patients exhibited a correlation between stromal and intraepithelial CD8^+^ T cells on day 10 post-treatment including polyfunctional CD8^+^PD1^+^GrB^+^ and CD8^+^PD1^+^GrB^+^Ki67^+^ (**Fig. 4H, I**) which was not present at baseline, nor was it observed in NR patients at any time point (**Fig. 4H**). These findings indicate that LDRT treatment induces an influx of polyfunctional CD8^+^ cells from the stroma into the tumor islets of R patients subsequently enabling tumor recognition and anti-tumor response.

Additionally, we observed a non-significant trend towards reduced NK cell numbers in Rs and an increase in NK cells in NRs following LDRT+/− Cy (**Supp. 5F**). We did not detect a significant change in the Foxp3^+^/CD8^+^ T cell ratio between response categories (**Supp. 5G**) nor between patients treated with or without low-dose Cy (**Supp. 5H**).

Overall, our data suggests that tumors with high DNA repair responses and HRD footprints retain a degree of immunogenicity which when mobilized by LDRT can promote tumor-infiltrating T cells functionality and enable adaptive responses to ICB therapy.

## DISCUSSION

This phase I trial evaluated the RACIN regimen in heavily pretreated patients with multimetastatic solid tumors characterized by CD8 immune-exclusion, PD-L1 negativity, and low TMB indicative of absence of mutations. LDRT was applied to all metastatic deposits to modulate the TME and enhance systemic immune responses. The primary outcomes included toxicities ≥ grade 3 that occurred in 33% of patients from Cohorts 1 and 2, consistent with prior ipilimumab and nivolumab studies (36), but persistent immune-related toxicities were attributed to the interaction of ipilimumab and metronomic Cy. A protocol amendment randomizing patients to either ipilimumab/nivolumab or Cy/nivolumab reduced toxicity rates to 12–21% without new safety concerns, although 60% of patients in Cohort 3 discontinued treatment, mainly due to rapid disease progression. Secondary outcomes of efficacy remained limited, with an ORR of 8% and a DCR of 42%. Among them, a patient with gallbladder cancer achieved a short-term PR and a patient with HGSOC who achieved CR remains disease-free two years after completing study treatment and is currently receiving Nivolumab off-trial for over a year. Importantly, our complete responder was allocated to cohort 3B and received ipilimumab combined with nivolumab.

Welsh et al. (37) conducted a phase II trial combining ipilimumab with HDRT and LDRT for metastatic liver and lung lesions, showing a median progression-free survival of 2.9 months and a 26% clinical benefit rate for nonirradiated tumor volumes. Notably, non-targeted lesions receiving LDRT were more likely to respond (31% vs 5%, p = 0.0091), highlighting the potential of LDRT to enhance systemic immune responses without increasing adverse events. In comparison, our study demonstrated a higher DCR of 42%, which may be attributed to the irradiation of all visible metastatic lesions combined with a more intensified systemic treatment regimen. Indeed, Schoenfeld et al. (38) treated PD-(L)1-resistant NSCLC with a combination of durvalumab and tremelimumab alongside either LDRT or HDRT. However, they irradiated only one or two lesions per patient and found no clinical benefit, likely due to insufficient immune stimulation resulting from limited irradiation and suboptimal timing relative to ICI. Similarly, Monjazeb et al. (39) tested LDRT and hypofractionated RT in combination with ICB in metastatic colorectal cancer and also observed no significant clinical benefit. This suggests that LDRT alone may be insufficient to overcome resistance in immunotherapy-refractory tumors unless combined with more comprehensive tumor irradiation or additional immune-modulating strategies. The failure of these trials underscores the complexity of translating preclinical immunoradiotherapy findings into clinical success and reinforces the need to refine radiation dose, fractionation, and sequencing to maximize immune activation.

Despite evidence that LDRT influenced the TME in our patients, the low immunogenicity, excluded immune phenotype and aggressive nature of these tumors limited clinical benefit. This challenging cohort, inherently unlikely to respond to ICB alone, underscored the difficulty of achieving durable outcomes. Based on these findings, exploratory aims of the study were to identify biomarkers for improved patient selection in future clinical trials.

We observed increased DNA damage response (DDR) signatures and mutations in genes mediating DNA damage repair responses in the malignant TME compartment of responder and metabolic responder patients. Importantly LDRT further exacerbated DNA damages *in situ* in lesions responding to LDRT. Notably our sole CR patient with HGSOC carried co-occuring mutations in *ATM*, *CDK12* and *TP53* and DNA footprints of HRD such as LST, and signature 3, all indicative of HRD. The insights provided by the analysis of our clinical cohort elucidate the molecular underpinnings of LDRT and ICB responsiveness. They suggest that response to LDRT and ICB could potentially be predicted by examining mutations or the transcriptomic profiles of the DNA damage and DNA repair pathways in immune-excluded tumors. They also indicate that mutations in these pathways could be sensitizing tissues to LDRT and therefore are important for combinatorial LDRT and ICB therapy. Notably, recent findings indicate that co-occurring mutations in HRD genes of primary HGSOCs predict long-term survival in HGSOCs (40). This suggests that tumors with defective DNA repair not only exhibit increased sensitivity to DNA-damaging therapies but may also create a more favorable immune microenvironment. In our study, LDRT exacerbated existing DNA damages *in situ* enhancing ICB efficacy, seeding the idea that DDR deficiencies and potentially inducers of DDR (i.e. those targeting the ATM, ATR, PARP etc. proteins) can sensitize tumors to LDRT and enable immunotherapy responses.

Our tumor cohorts, immune excluded by design, presented a limited presence of CD8^+^PD1^+^ cells in the tumor epithelium at baseline and hence not expected to respond to ICB. These immune-excluded tumor atlases were marked by a complete absence of DC2 and immunoreactive CXCL9/10^+^ and ISG^+^ myeloid cells, reflecting a highly immune suppressive TME, despite pre-existing immunogenicity due to loss of DDR. The low presence of proliferative T cells, especially in more proliferative tumors, further underscored the challenge in achieving immune activation. In our cohort these tumors were predominantly infiltrated by MAIT CD8 TILs and immature myeloid cells which can contribute to a more locally immune-suppressed TME and underscore the need to re-activate local myeloid populations (41).

Despite the odds, our findings revealed that even a small number of CD8^+^PD1^+^/exhausted T cells, found to be in contact with myeloid niches, can be sufficient to induce a tumor response, and that the viability of these cells is not compromised by LDRT. Indeed, the presence of exhausted CD8^+^ TILs in patients showing local tumor response suggests pre-existing tumor immune recognition through chronic antigen exposure, also known for responsiveness to ICB in non-small cell lung cancer (NSCLC) and melanoma (42–44). LDRT delivery increased the trafficking of excluded CD8 TILs into the tumor epithelium and reinvigorated their functionality, marked by higher expression of Ki67, GrB, and/or PD1 in responder patients. This phenomenon was not observed in non-responders whose CD8 TIL states displayed increased radiosenstivity. Although the impact of hypofractionated RT on IFN-I mediated activation of myeloid cells and its effect on T cell reinvigoration has been previously documented (14, 45, 46), this phase I study is the first to report LDI-induced changes in CD8^+^ T cells within the TME of cancer patients with immune excluded tumors who subsequently responded to ICB. Consistent with prior studies using 20 Gy RT (35), we found that LDRT did not induce T cell apoptosis in the few patients that we could analyze, even with large irradiated fields. Remarkably, patients who showed an increase in intraepithelial CD8^+^ TILs after LDRT were more likely to carry mutations in DNA repair and cell cycle related genes, underscoring that tumor cells with impaired DDR may be more susceptible to LDRT, potentially enhancing immunogenic cell death and subsequently boosting CD8^+^ T cell infiltration and response to ICB.

Our findings in heavily pretreated patients with multimetastatic solid tumors align with other studies suggesting the potential for LDRT to enhance immune responses. A phase 2 trial in metastatic patients refractory to ICB demonstrated increased infiltration of CD4^+^, CD8^+^, and NK cells in the TME after HDRT and LDRT (47). Similarly, in a recent phase II study by Altorki et al. (48), patients with early-stage NSCLC treated with neoadjuvant durvalumab and 3 × 8 Gy stereotactic HDRT showed a systemic adaptive immune response, evidenced by increased intra-tumoral CD103^+^ T cells and immune-related gene expression. Notably, pre-treatment levels of circulating CD103^+^ T cells correlated with major pathological response (MPR), highlighting their predictive value. A phase I trial in NSCLC found that concurrent HDRT and ICB improved responses, especially in high-aneuploidy, immune-cold tumors, which had less progression at unirradiated sites (49). Indeed, recent studies show that tumor aneuploidy can impair anti-tumor immunity through mechanisms like PD-L1 downregulation (50) and suppression of intratumoral CD8^+^ T cells (49), contributing to immune evasion by reducing T cell infiltration and dampening immune activation. However, chromosomal instability-driven tumors depend on cGAS-STING and IL-6-STAT3 signaling for survival (51), creating a potential vulnerability. Notably, IL-6R inhibition with tocilizumab selectively impaired chromosomal instability-high tumors, suggesting that targeting inflammatory pathways could enhance LDRT-ICB responses. These findings suggest that aneuploidy can promote immune evasion but also pose a vulnerability for combined therapies like radiation and ICB. In cancers with low TMB, and immune-excluded/cold tumors aneuploidy, DDR signatures and/or mutations could constitute biomarkers for patient selection and stratification in future clinical trials of LDRT and immunotherapy.

While our study and the aforementioned trials focus on tumor-directed LDRT, an alternative strategy, iLDRT, aims to enhance systemic immunity via gut microbiota modulation. A Phase 2 trial testing iLDRT with anti-PD-L1 therapy suggests it may improve responses in patients progressing on immunotherapy, without direct tumor irradiation (20). Unlike tumor-directed RT, which boosts local TIL infiltration, iLDRT activates systemic immunity via gut-immune crosstalk. While distinct in mechanism, it may complement tumor-directed LDRT, warranting future studies to compare immune pathways, biomarkers, and potential synergy.

Our study has several limitations, including limited clinical activity with the LDRT-ICB combination, leading to early trial termination. We lacked sufficient patient numbers to assess the clinical activity of 0.5 Gy versus 1 Gy, and paired samples were scarce for the 0.5 Gy cohort. Despite those, we observed LDRT-induced changes in the TME, highlighting the potential for further research. Sample availability and tumor heterogeneity also impacted our analysis, but we successfully constructed a TME atlas for immune-excluded tumors and evaluated LDRT effects.

In summary in this pre-selected population, we identified a subgroup of immune-excluded patients with DNA repair signatures and exhausted CD8^+^ T cells that could be reinvigorated by LDRT in responder patients. These patients may potentially benefit from LDRT and ICB serving as biomarkers for selecting patients in future clinical trials of radiation and immunotherapy.

## Data Availability

All data produced in the present study are available upon reasonable request to the authors.

## Acknowledgements

We are grateful to the patients and their families for their dedicated collaboration.

We thank Jean-Paul Rivals, Lana E. Kandalaft.and all the team from CHUV Biobank - Center of Experimental Therapeutics (CTE), CHUV; Nathalie Piazzon from the Institute of Pathology, Lausanne University Hospital for their assistance. We thank the Lausanne Genomic Technologies Facility for RNA-seq analysis.

## Funding resource

This work was supported by the Ludwig Institute for Cancer Research, and grants from Bristol-Myers Squibb, the Prostate Cancer Foundation Challenge Award (18CHAL08), and the Cancera, Biltema and Paul Matson Foundations. This Research Project was partially supported by the Spanish Society of Medical Oncology (SEOM).

