## Supplemental data for "Combinatorial delivery of low-dose irradiation and immunotherapy to patients with immune-excluded tumors effectively enhances CD8^+^ T cell functionality"

**Supplementary Material:**

- **Materials and Methods**
- **Supplementary Figures 1-5**
- **Supplementary Tables 1-5**
- **Supplementary References**

### MATERIALS AND METHODS

#### Study design:

The primary objective of the study was to determine the safety and tolerability of the combination of the backbone treatment, comprising nivolumab, ipilimumab, aspirin/celecoxib, and metronomic Cy in association with escalated LDRT and to determine the maximum tolerated dose (MTD) of RT. DLTs were defined as toxicities related to LDRT, and backbone limiting toxicities (BLTs) were defined as toxicities related to the backbone combination. Toxicity was measured using the Common Terminology Criteria for Adverse Events (CTCAE) v.4. Supp. Table 5 outlines the definitions of DLTs and BLTs, specifying their assignment to individual treatments or the combination. Both DLTs and BLTs were defined as such if they occurred between Cycle 1 Day 1 and Cycle 2 Day 1 (pre-dosing, 8 weeks) during the combined treatment regimen. This regimen included LDRT at either 0.5 Gy or 1 Gy per fraction every two weeks (total dose: 6 Gy or 13 Gy), metronomic Cy (200 mg/m² every two weeks), anti-PD1 (nivolumab, 240 mg every two weeks), and anti-CTLA4 (ipilimumab, 1 mg/kg every six weeks). Cycle 0 involved the administration of LDRT ± metronomic Cy, aimed at reprogramming the TME to enhance the efficacy of the subsequent full combination therapy; cycle 0 was not assessed for DLT/BLT period. Following four cycles of the combinatorial treatment, patients achieving CR, PR, or SD, who tolerated the treatment without significant side effects, were permitted to continue anti-PD1 therapy for up to one year.

<u>Cohort 1-2 (Fig. 1A)</u>

Cohort 1 followed a 3+3 algorithm adapted for multidrug combination therapy. For cohort 2, a fixed number of 6 patients per arm were included, using also an algorithm adapted to multidrug combination. Patients included in cohort 1 and 2 received LDRT with 0.5 or 1 Gy per fraction respectively. LDRT was delivered every two weeks, for a total dose of 6 Gy or 12 Gy respectively and was delivered to  target and non-target metastatic lesions in combination with low-dose Cy (200 mg/m2 Q2W), Nivolumab (anti-PD1) 240 mg Q2W and Ipilimumab (anti-CTLA4) 1 mg/kg Q6W, for up to 24 weeks, and Aspirin (300 mg p.o, QD). After four cycles of Ipilimumab/Nivolumab, eligible patients which were not in progression by RECIST/PCWG3 could continue Nivolumab maintenance (Q2W) with Aspirin (QD) until disease progression or toxicity. All metastatic lesions were irradiated when possible while preserving the bone marrow as much as possible.

Cohort 3, arm A and B (Supp. 1A, B)

Considering reported toxicity in cohorts 1 and 2, the investigators proposed a protocol amendment to adjust the treatment regimen. Specifically, it was decided to avoid the simultaneous delivery of Ipilimumab and Cyclophosphamide in the same patient to prevent excessive Treg depletion (1-3), which could result in insufficient T helper and cytotoxic T cell suppression (4), hence leading to excessive auto-reactive immune cells.

Cohort 3 had a randomized, non-comparative design to determine a safe backbone combination. Patients were randomized into two arms (A and B), with six 6 patients per arm. All patients received radiation (1Gy in both arms) and celecoxib (200mg p.o., twice daily) as well as Nivolumab combined with either low-dose Cy (Arm A) or Ipilimumab (Arm B). The safety of each arm was evaluated separately (Supp. 1A, B). Doses and frequency of drug delivery was the same as in cohorts 1 and 2. Aspirin was substituted by celecoxib due to lack of availability of ranitidine (H2 antagonist required as gastric protector when administering continuous aspirin) in the Swiss market.

Ipilimumab (anti-CTLA-4) and Nivolumab (anti-PD-1) were chosen as ICB based on preclinical data which showed that 1 Gy induced upregulation of co-inhibitory molecules PDCD1 (PD-L1) and CTLA-4 (5). Aspirin/Celecoxib were chosen as a cyclo-oxygenase 1 and 2 (COX1/2) inhibitors, with the goal of decreasing production of prostaglandins (6). The presence of prostaglandin E2 has been associated with unfavorable prognosis (7), as the COX2-

PGE2 axis has a pro-tumoral effect that contributes to tumor cell proliferation and differentiation, angiogenesis, and tumor immunosuppression (7).

All patients provided written informed consent before enrollment. The study was conducted in accordance with ethical principles founded in the Declaration of Helsinki. The trial was approved by the Institutional Review Board of the Centre Hospitalier Universitaire Vaudois (CHUV), Lausanne, Switzerland.

**Study participants/Patients**

Eligible patients were adults with histologically proven solid tumors who had progressed to at least one standard therapy for advanced disease. Patients were required to have a good general health status (ECOG PS 0-1). Treatment with previous anti-PD(L)1 or anti-CTLA4 was allowed. A pre-screening biopsy was mandatory for inclusion, and patients required to have absence of tumor-infiltrating intraepithelial $CD8^+$ T cells by IHC defined as $< 5$ $CD8^+$ cells per high power field of tumor, analysed by a certified pathologist. Patients were required to have at least one lesion with measurable disease (except for prostate cancer patients) as defined by RECIST v1.1 criteria for response assessment. Participants with lesions in a previously irradiated field as the sole site of measurable disease were permitted to enrol provided that the lesion(s) had demonstrated clear progression prior to inclusion. Patients were required to have adequate normal organ and marrow function, defined as haemoglobin $\geq 90$ g/L; absolute neutrophil count $\geq 1.5$ G/L; platelet count $\geq$100G/L; serum creatinine $\leq 1.5$ x ULN; and AST and ALT $\leq 2.5$ x ULN. Patients with autoimmune diseases were excluded, as well as patients with symptomatic brain metastases unless stable for 6 months. Data cut-off for the final analysis was performed on 11 December 2023.

**Outcomes**

Clinical data were captured electronically using the iMedidata database up to 11 December 2023,, which was the cut-off date for final analysis. The safety and tolerability of the trial treatment was evaluated by the occurrence of toxicities and adverse events initiated in the DLT/BLT period (as well as in the overall period). Toxicity was assessed in the safety population (patients who received at least one dose of trial treatment). Classification of severity and causality was performed according to CTCAE v4.03. Best overall response and objective response (CR or PR) rate (ORR) by RECIST 1.1 (and by PCWG3 for prostate cancer) were presented in the efficacy population (all patients with measurable disease at enrolment who received at least one dose of trial treatment), by cohort/arm. Information was also provided for irradiated lesions only. Disease control (CR, PR or SD) rate (DCR) at 3, 6 and 12 months by RECIST 1.1 (and by PCWG3) was analysed by cohort/arm. Time to progression (TTP) was evaluated in the efficacy population. Overall survival (OS) and progression-free survival (PFS) were calculated from the enrolment date in the efficacy population. 18F-FDG-PET/CT was performed in seven patients, while 68Ga-PSMA-PET/CT was performed in five PCa patients.

**Immunohistochemistry, TIL assessment and scoring**

Biopsies were screened to assess tumor viability and quantify the intra-tumoral $CD3^+$ and $CD8^+$ infiltration by a dedicated Pathologist. For each case, representative tumor slides with the most viable component and higher inflammatory infiltrate were selected for immunohistochemical (IHC) staining on formalin-fixed paraffin-embedded (FFPE) whole tissue sections. CD3 (2GV6, CONFIRM, Rabbit Monoclonal, Roche, Basel, Switzerland) and CD8 (C8/144B, Mouse Monoclonal, Dako, Glostrup, Denmark) expressions were assessed. Briefly, immunohistochemical staining (IHC) was performed using the Ventana Benchmark Ultra (Roche Ventana Tucson, Arizona, USA) by following the manufacturer's instructions. Four μm-thick FFPE sections were subjected to routine deparaffinization, rehydration, and antigen retrieval procedures. Each section was incubated with the primary antibody, followed by the secondary antibody. The ultraView Universal DAB Detection Kit (Ref: 05269806001 , Roche Ventana) were used as detection system. Tissue counterstaining was performed with Hematoxylin from Gil II solution (Ref: 105175, MERCK). Sections of human tonsil were used as positive control. Evaluation was performed independently by one pathologist (JD) without knowledge of clinical information. For each sample, at least 10 HPF whenever feasible with a diameter of 500 μm were selected. These fields were distributed over the whole tumor sample. Intra-tumoral T lymphocytes were qualitatively assessed by CD3 (low to high), along with spatial distribution (stroma vs. tumor) and heterogeneity. CD8/CD3 ratio and CD8 ranges per HPF were then evaluated. The final TILs score was the mean intra-tumoral $CD8^+$ cells in at least 10 HPF. The tumor-infiltrating $CD8^+$ T-cells were evaluated and classified as "intra-tumoral" if they were in direct contact with tumor cells. Cells stained positive in the stromal compartment and within the borders of the invasive tumor or in areas of necrosis were not evaluated. For cases with significant region variations in lymphocyte distribution, each region was evaluated separately, and an average value was assessed in > 10 HPF whenever possible.

**Multispectral immunofluorescence tissue staining and image analyses**

Tissue quality control was performed prior to multiplex immunofluorescence analysis using corresponding HE stains. Tumor content was assessed including areas of viable tumor cells, necrosis, inflammation.

Multiplexed staining was performed on 4-micrometer formalin-fixed paraffin-embedded (FFPE) tissue sections on automated Ventana Discovery Ultra staining module (Ventana, Roche). Slides were placed on the staining module for deparaffinization, epitope retrieval (64 minutes at 98°C) and endogenous peroxidase quenching (Discovery Inhibitor, 8 minutes, Ventana).

Multiplex staining consists in multiple rounds of staining. Each round includes non-specific sites blocking (Discovery Goat IgG and Discovery Inhibitor, Ventana), primary antibody incubation, secondary HRP-labeled antibody incubation for 16 minutes (Discovery OmniMap anti-rabbit HRP (Ventana, # 760-4311) or anti-mouse HRP (Ventana, #760-4310)), OPAL™ reactive fluorophore detection (Akoya Biosciences, Marlborough, MS, USA) that covalently label the primary epitope (incubation : 12 minutes) and then antibodies heat denaturation. Nuclei were visualized by a final incubation with Spectral DAPI (1/10, FP1490, Akoya Biosciences) for 12 minutes.

The first panel was optimized to look at the T-cells activation. Sequence of antibodies used in the multiplex with the associated OPAL are the following : 1$^{st}$: mouse monoclonal anti-human PD1 antibody (2µg/mL, NAT105, Biocare, 1hour, RT), OPAL570 ; 2$^{d}$ : rabbit anti-CD3 antibody (1.5µg/mL, DAKO, 1hour, 37°C), OPAL480; 3$^{rd}$ : mouse monoclonal anti-human Granzyme B antibody (1 µg/ml GrB-7, Monosan, 1hour, RT), OPAL620 ; 4$^{th}$: rabbit anti-human Ki67 antibody (0.06 µg/ml, SP6, Spring, 1hour, RT), OPAL520; 5$^{th}$ : mouse anti-pan-Cytokeratin antibody (1 µg/ml, Clone AE1/AE3, Dako, 1hour, 37°C), OPAL690; 6$^{th}$ : rabbit anti-CD8 antibody (0.3 µg/mL, clone SP16, Cellmarque, 1hour, 37°C), OPAL780.

The second panel was optimized to look at the T-cells subset. Sequence of antibodies used in the multiplex with the associated OPAL are the following : 1$^{st}$: rabbit anti-CD4 antibody (0.3 µg/mL, SP35, Cellmarque, 1hour, 37), OPAL570 ; 2$^{d}$ : rabbit anti-FOXP3 antibody (0.06 µg/mL, SP16, ABCAM, 1hour, 37°C), OPAL520; 3$^{rd}$ : rabbit monoclonal anti-human CD56 antibody (0.06 µg/ml, MRQ-42, Cellmarque, 1hour, RT), OPAL620 ; 4$^{th}$: rabbit anti-CD3 antibody (1.5µg/mL, DAKO, 1hour, 37°C), OPAL480; 5$^{th}$ : mouse anti-pan-Cytokeratin antibody (1 µg/ml, Clone AE1/AE3, Dako, 1hour, 37°C), OPAL690; 6th : rabbit anti-CD8 antibody (0.3 µg/mL, clone SP16, Cellmarque, 1hour, 37°C), OPAL780.

The third panel was optimized to look at DNA damage. Sequence of antibodies used in the multiplex with the associated OPAL are the following : 1st: rabbit anti-pSTING antibody (1/50, E9A9K, BIOCONCEPT, 1hour, RT), OPAL520 ; 2d : mouse anti-gH2Ax antibody (0.5 µg/mL, JBW301, Millipore, 1hour, RT), OPAL570; 3rd : rabbit anti-pSTAT antibody (1/100, 58D6, Cellsignaling, 1hour, RT), OPAL620 ; 4th: mouse anti-CD11c antibody (1/100, 5D11, Cellmarque, 1hour, 37°C), OPAL690; 5th : mouse anti-pan-Cytokeratin antibody (1 µg/ml, Clone AE1/AE3, Dako, 1hour, 37°C), OPAL480; 6th : rabbit anti-CD8 antibody (0.3 µg/mL, clone SP16, Cellmarque, 1hour, 37°C), OPAL780.

The fourth panel was optimized to look at DC-Kissing/B-cell. Sequence of antibodies used in the multiplex with the associated OPAL are the following : 1st: mouse monoclonal anti-human PD1 antibody (2µg/mL, NAT105, Biocare, 1hour, RT), OPAL570 ; 2d : rabbit anti-CD19 antibody (1/50, BT51E, Novacastra, 1hour, RT), OPAL620; 3rd : mouse monoclonal anti-CD68 antibody (0.06 µg/ml, PG-M1, Dako, 1hour, 37°C), OPAL480 ; 4th: mouse anti-CD11c antibody (1.5µg/mL, 5D11, Cellmarque, 1hour, 37°C), OPAL520; 5th : mouse anti-pan-Cytokeratin antibody (1 µg/ml, Clone AE1/AE3, Dako, 1hour, 37°C), OPAL690; 6th : rabbit anti-CD8 antibody (0.3 µg/mL, clone SP16, Cellmarque, 1hour, 37°C), OPAL780.

Multiplex IF images were acquired on PhenoImager allowing a whole slide multispectral imaging acquisition (Akoya Biosciences). IF signal extractions from qptiff were performed using our in-house developed software IFQuant (https://github.com/BICC-UNIL-EPFL/IFQuant), enabling automatic tissue segmentation into stroma and tumor (intraepithelium) based on CK and DAPI staining and per-cell analysis of IF markers of multiplex stained tissue sections. Final output consists in complete tsv file with raw data with cell coordinates, localization and phenotyping, and excel file with calculated densities (cell/mm$^2$) of each phenotypes using IFQuant-integrated R-script.

**Bulk RNA processing**

Total RNA was extracted from snap frozen biopsies, two time points were analyzed (baseline and day 10). Directly after the thawing, samples were mechanically lyzed with a metallic bead in TRIzol solution using the Tissue Lyzer (InVitrogen). After a phase separation with chloroform, RNA purification was done with the RNeasy mini or RNeasy micro kit (Qiagen) depending of the size of input material. The quality of resulting RNA was evaluated by a Nanodrop spectrophotometer (Thermofisher) and a Fragment Analyzer (Agilent). Quantification with Qubit HS RNA assay kit (InVitrogen) was also done. Sequencing libraries were constructed according to *Illumina* protocol using the Illumina TruSeq Stranded Total RNA kit and sequenced on a Novaseq sequencer (Lausanne Genomic Technologies Facility) according to manufacturer recommendations.

**Bulk RNA Sequencing**

RNA quality was assessed on a Fragment Analyzer (Agilent Technologies) and the RNAs had RQNs from 6.1 - 9.9. RNA-seq libraries were prepared from 100 ng of total RNA with the Illumina Stranded Total RNA Prep with Ribo-Zero Plus reagents (Illumina) using a unique dual indexing strategy and following the official protocol. Libraries were quantified by a fluorimetric method (QubIT, Life Technologies) and their quality assessed on a Fragment Analyzer (Agilent Technologies). Sequencing was performed on an Illumina NovaSeq 6000 for 300 cycles (paired-end 150 reads). Sequencing data were demultiplexed using the bcl2fastq2 Conversion Software (version 2.20, Illumina).

Two samples had low RNA quantity and were sequenced separately. RNA-seq libraries were prepared by first generating double-stranded cDNA with the Ovation RNA-Seq System V2 (TECAN). 100 ng of the resulting double-stranded cDNA were used for sequencing library preparation with the Illumina DNA Prep kit (Illumina) according to the protocol supplied by the manufacturer. Sequencing was performed on an Illumina NovaSeq 6000 for 300 cycles (paired-end 150 reads). Sequencing data were demultiplexed using the bcl2fastq2 Conversion Software (version 2.20, Illumina).

**Bulk RNA-seq Alignment, Processing and Sample Selection**

The bulk RNA-seq data was aligned and sorted against GRCh37 reference genome, using Samtools (version 1.8) and STAR Aligner (version 2.6.0) software, following the methodology presented in Barras and colleagues (ATATIL). Quality control (QC) metrics for the data were generated using FastQC (version 0.11.7). Finally, Htseq (version 0.9.1) processed the filtered data returning the raw count matrix used for downstream analysis.

The raw counts were processed using edgeR R package (version 3.34.1). The data was filtered based on expression by using the function "filterByExpr", followed by normalization using "calcNormFactors" applying the TMM method as proposed by Robinson and Oshlack (8). As the data was processed in two batches, "ComBat" function from the sva package (version 3.44.0) was employed to reduce the introduction of variation in the dataset by using the cohorts as covariate.

Principal Component Analysis (PCA) highlighted tumor type and biopsy location to be major sources of variance in the data. To gain in statistical power, the analysis was focused on groups with at least three samples sharing tumor type. This condition was filled for samples with ovarian, prostate and colon as their primary malignancies. The colon cancer samples were removed due to the lack of response in this subgroup which hindered the comparison based on responder status.

**Tissue processing for scRNA**

The day of the assay, frozen tissues were thawed in 10ml of R10 before cutting and fresh biopsies were chopped upon reception. Some biopsies were processed fresh. Tissues were cut with scalpels in a Petri dish containing few microliters of R10 medium. Fragments were transferred in a 2ml Eppendorf tube with 1ml of digestion mix (RPMI + 2% Gelatin + 50 IU/mL Collagenase I + 50 IU/mL Collagenase IV + 30 IU/mL Deoxyribonuclease I + 0.1% RNasin and incubated 15-20 minutes at 37°C under agitation at 130 rpm. Digestion reaction was stopped by adding 500ul of R10 to the mix. Digested fragments were filtered on a 70um strainer placed on the top of a 50ml Falcon tube. 1ml of R10 was added to the filter before filtering the cell suspension. The remaining pieces were smashed against the filter using the plunger of a 2ml syringe and washed with 10-15ml of R10. Suspension was centrifuged 5min at 400g and supernatant discarded. For counting and estimation of viability with Trypan blue, cells were resuspended in an appropriate volume (300 to 500ul) of PBS + 1%Gelatin + 0.1%RNasin according to the pellet size.

In order to preserve cell viability, only ¼ of cells were stained for CD45+ and EpCAM+ population. The rest of the sample was kept on ice. Cells were FcR blocked in 50ul of 1xPBS + 1%FBS + 0.1%RNAsin for 15min at RT. Then, they were directly stained with CD45-FITC and EpCam-PE for 20min at 4°C. After a wash with PBS + 1%FBS + 0.1%RNAsin cells were resupended in 500ul of sorting solution (PBS + BSA0.04% + RNAsin 0.1%). Then, viability stain was performed with RedDot1 (1/200) for 10min at 37°C. DAPI (1/1000) was added just few minutes before the sort.

*FACS sorting of cells*

$CD45^+$ and $EpCam^+$ population frequencies were estimated by FACS. Then all viable cells were sorted in a 0.2ml PCR tube on a MoFlo Astrios (Beckman Coulter) containing 10ul of PBS + 0.04% BSA + 0.1% RNasin. After sort, if count was > 30’000, cell number was manually estimated with an hemacytometer and viability was assessed using Trypan blue exclusion. If FACS count was < 20’000, cells were encapsulated without prior manual counting.

*Encapsulation and library construction*

When cell number was sufficient, a maximum of 15’000 live cells per sample was loaded into the Chromium machine for encapsulation and barcoding using the Next GEM Single Cell 5’ Library and Gel beads kit v1.1. Resulting cDNA was amplified with 15 PCR cycles then 5GEX and V(D)J libraries were constructed according to *10XGenomics* protocol (CG000208). Quality controls were performed on a Fragment Analyzer (Agilent) and quantification of libraries was done with the Qubit HS dsDNA assay kit (InVitrogen).

*Sequencing*

5GEX and V(D)J libraries were sequenced on an Illumina HiSeq4000 or a NovaSeq sequencer following *10XGenomics* parameters. The median depth per cell for sequencing was 20’000 reads for 5GEX libraries and 5’000 reads for V(D)J libraries.

**scRNA sequencing**

Sequencing was performed with Illumina technology either with the HiSeq4000 or the NovaSeq6000 with 28 cycles read1, 10 cycles i7 index read, 10 cycles i5 index read, and 91 cycles read2. For HiSeq4000, cluster generation was performed with 1.25 nM library pool spiked with 10% PhiX, using the Illumina HiSeq 3000/4000 PE Cluster Kit reagents and sequencing with the HiSeq 3000/4000 SBS Kit reagents. For NovaSeq6000, cluster generation was performed with 0.6 nM library pool spiked with 1% PhiX, using the NovaSeq 6000 cluster cartridge v1.5 and sequencing with the NovaSeq 6000 SBS cartridge 100 cycles v1.5. In all cases, the actual amount of individual libraries multiplexed in the sequencing pools was adjusted based on the number of targeted sequencing reads. Sequencing data were demultiplexed using the bcl2fastq2 Conversion Software (v. 2.20, Illumina).

**scRNA-seq Processing, Integration and Dimensional reduction**

The scRNA-seq reads were aligned to the GRCh38 reference genome and quantified using Cell Ranger count (10X Genomics, version 4.0.0). We only retained gene-code matrices with unique molecular identifier (UMI) counts who passed the cell detection threshold. Further processing and analysis of the data was performed using the R package Seurat (version 4.3.0) using default parameters unless stated otherwise.

Initially, a lenient approach to QC was applied to keep a good representation of the malignant compartment, potentially treatment-induced damaged cells, and doublets. Low-quality cells and doublets were successfully identified but removed from subsequent downstream analysis. The dataset consisted of 47’453 cells. These were then normalized by total counts and log-transformed using “NormalizeData” function from Seurat. The normalized data was scaled using the “ScaleData” function.

To address the potential confounding impact of batch effect on the transcriptional data, the samples were integrated using the workflow described by Satija Lab (https://satijalab.org/seurat/articles/integration_introduction.html). This approach involved identifying pairs of cells from different samples which are mutual nearest neighbors, which share sources of variation and are used as anchors for the integration. We then applied the “ScaleData” function to calculate z-scores for each gene and performed PCA on the scaled integrated data. We further applied Uniform Manifold Approximation and Projection (UMAP) using the first 20 PC dimensions to reduce the dimensionality of the data and plot the projection of the whole dataset.

*scRNA-seq Immune Compartment Annotation:*

We used SingleR (version 1.10.0) and the signatures provided by Blueprint and ENCODE, consisting of 259 bulk RNA-seq pure samples, and accessible in the celldex package (version 1.6.1). We initially used this annotation to separate de dataset into the following major populations: CD8, CD4, Myeloid, Natural killer (NK) cells, B-cells, and malignant cells.

Quality control (QC) was applied to the immune-specific datasets. Filtering was performed using mitochondrial and ribosomal RNA as QC metric with cells having expression levels above 20 and 60 percent, respectively, being deemed of low quality and filtered out. This resulted in the retention of 30’029 $CD45^+$ cells with an average of 1’375 gene expressed per cell and averaged unique number of transcripts of 3’643.

The cell-specific data subsets, with a minimum of 30 cells, were integrated and scaled as previously described. Additionally, for the $CD4^+$ and $CD8^+$ specific samples, we regressed the TCR-alpha and TCR-beta genes while scaling the data to avoid clonotype-specific skewing of the data. The 1’000 most variable features were identified using “FindVariableFeatures”. The k-nearest neighbors for each cell were computed using the function “FindNeighbors” based on the UMAP reduction. This was followed by data clustering for different resolutions (0.1 to 2 by 0.1 increments) by using “FindCluster”. Markers for each cluster were identified using the “FindAllMarkers” function on the RNA assay and using a Wilcoxon Rank Sum test.

Cycling cells were identified by their G2/M phase and S phase score which was computed using “CellCycleScoring” on the different datasets with the cell-cycle genes provided by Seurat. To further refine the annotation of immune cell states in the TME, we applied the same strategy as presented by Barras and colleagues(9). We identified a few clusters of cells in different population that stood out by their low Pearson correlation scores with any cell state signatures provided by Barras and colleagues. These were subsequently labelled by using literature-based markers from following sources: Zheng et al. (10), which led to the annotation of $CD8^+$ MAIT, $CD4^+$ MAIT cells and $CD4^+$ Type 1 IFN. The cytotoxic NK and regulatory NK cells were annotated based on markers reported by Cao et al (11).

*Quantification and Pathway Enrichment Analysis*

The quantification of immune cells was calculated out of all $CD45^+$ cell proportion for each sample. To perform the pathway enrichment analysis, signature scores were measured using single sample gene set enrichment analysis (ssGSEA) as proposed by the GSVA R package (version 1.44.5). The gene components for the pathways were taken from the Molecular Signatures Database (MSigDB) (https://www.gsea-msigdb.org/gsea/msigdb/), specifically from the H (hallmark) and C2 (curated gene) collections.

To visualize the results, we generated violin-, line- and bar-plots using the R package ggplot2 (version 3.3.6). The heatmaps were created using the function dedicated to this in the heatmap (version 1.0.12) R package, plotting the z-scores for each pathway.

For both the BulkRNA and scRNA data was analyzed using R software (version 4.2.1) unless stated otherwise.

#### Data availability

Bulk-RNA and single-cell RNA sequencing data will be made publicly available in the Gene Expression Omnibus (GEO) at the time of publication.

**Targeted high-throughput DNA sequencing (400-gene panel)**

Targeted high-throughput sequencing (HTS) was performed in parallel on DNA extracted from FFPE tumor tissue sections at baseline (Maxwell 16 FFPE Plus LEV DNA Purification kit, Promega, Madison, WI) and matched constitutional DNA extracted from blood (Maxwell 16 LEV Blood DNA kit, Promega), using a customized panel covering the full coding sequences of approximately 400 cancer-related genes, as follows:

Version 1 (394 genes): *ABL1 ABRAXAS ACVR1 ACVR1B ACVR2A AKT1 AKT2 AKT3 ALK AMER1 ANKRD11 APC APCDD1 AR ARAF ARID1A ARID1B ARID2 ARID5B ASXL1 ASXL2 ATM ATR ATRX AURKA AURKB AXIN1 AXIN2 AXL B2M BAP1 BARD1 BCL2 BCL2L1 BCL6 BCOR BCORL1 BIRC3 BLM BRAF BRCA1 BRCA2 BRD4 BRIP1 BTK CARD11 CASP8 CBFB CBL CCN6 CCND1 CCND2 CCND3 CCNE1 CD274 CDC73 CDH1 CDK12 CDK4 CDK6 CDK8 CDKN1A CDKN1B CDKN2A CDKN2B CDKN2C CEBPA CENPA CHD4 CHEK1 CHEK2 CHUK CIC CREBBP CRKL CTCF CTNNB1 CUL3 CUL4A CUL4B DAXX DCUN1D1 DDR2 DDX3X DICER1 DNMT1 DNMT3A DNMT3B DOT1L E2F3 EED EGFR EIF1AX ELOC EP300 EPHA3 EPHA6 ERBB2 ERBB3 ERBB4 ERCC2 ERCC3 ERCC4 ERG ERRFI1 ESR1 ETS2 ETV6 EWSR1 EZH2 FANCA FANCC FANCD2 FANCE FANCF FANCG FANCI FANCL FANCM FAT1 FAT3 FBXW7 FCGR2A FGF12 FGF19 FGF3 FGF4 FGFR1 FGFR2 FGFR3 FGFR4 FLCN FLT1 FLT3 FLT4 FNTB FOXA1 FOXL2 FOXP1 FRK FUBP1 FYN GATA1 GATA2 GATA3 GLI1 GNA11 GNAQ GNAS GPS2 GRIN2A GSK3B GSTP1 GUCY1A2 H3-3A H3-3B H3C2 HDAC9 HGF HLA-A HNF1A HRAS HSP90AA1 IDH1 IDH2 IGF1 IGF1R IGF2 IGF2R IKBKE IKZF1 IL6ST IL7R INPP4A INPP4B INSR IRS1 IRS2 JAK1 JAK2 JAK3 JUN KAT6A KDM5A KDM5C KDM6A KDR KEAP1 KIT KLF4 KMT2A KMT2C KMT2D KRAS LATS1 LATS2 LRP1B LRP6 LTBP4 MAP2K1 MAP2K2 MAP2K4 MAP3K1 MAP3K11 MAP3K13 MAP3K4 MAP3K8 MAPK1 MAPK3 MAX MCL1 MDC1 MDM2 MDM4 MED12 MEN1 MET MGA MITF MLH1 MRE11 MSH2 MSH6 MST1R MTHFR MTOR MUTYH MYC*

*MYCL MYCN MYD88 NBN NCOA3 NCOR1 NCOR2 NF1 NF2 NFE2L2 NFKBIA NKX2-1 NOTCH1 NOTCH2 NOTCH3 NOTCH4 NPM1 NQO1 NRAS NSD1 NTRK1 NTRK2 NTRK3 NUTM1 PAK1 PAK5 PALB2 PARP1 PARP2 PARP3 PARP4 PAX8 PBRM1 PCSK6 PDGFRA PDGFRB PDK1 PDPK1 PHLPP2 PIK3C2G PIK3C3 PIK3CA PIK3CB PIK3CD PIK3CG PIK3R1 PIK3R2 PIK3R3 PIM1 PLCG1 PLK2 PMAIP1 PMS1 PMS2 PNRC1 POLE POLR2A PPM1D PPP2R1A PPP6C PRKAR1A PRKDC PRSS1 PSMA6 PTCH1 PTCH2 PTEN PTPN11 PTPRD PTPRF PTPRS PTPRT PTPRU RAB35 RAC1 RAD21 RAD50 RAD51 RAD51B RAD51C RAD51D RAD52 RAD54L RAF1 RARA RASA1 RB1 RBM10 RET RHEB RHOA RICTOR RIT1 RNF43 ROS1 RPA1 RPS6KB2 RPTOR RSPO2 RUNX1 RUNX1T1 RYBP SDHB SDHD SETD2 SF3B1 SHQ1 SMAD2 SMAD3 SMAD4 SMARCA4 SMARCB1 SMARCD1 SMO SNX31 SOS1 SOX2 SOX9 SPEN SPOP SRC STAG2 STAT3 STAT5A STK11 STK19 STK40 SUFU SUZ12 SYK TACC1 TAOK1 TAOK2 TBX3 TERT TET1 TET2 TGFBR1 TGFBR2 TIPARP TMPRSS2 TNFAIP3 TNFRSF14 TNK2 TNKS TNKS2 TOP1 TP53 TP63 TRAF7 TRRAP TSC1 TSC2 TSHR VHL WEE1 WNT5A WT1 XPO1 XRCC1 XRCC2 XRCC3 ZBTB16 ZFHX3 ZNF703 ZNRF3.*

Version 2 (422 genes): genes included in version 1, from which were removed *ETV6 NUTM1 TMPRSS2*, and to which were added *BMPR1A CALR CSF1R CSF3R CYLD CYSLTR2 DDR1 EPHB1 ERCC1 FAS FH H3C3 HDAC2 LYN MLH3 MSH3 NKX3-1 PDCD1 PDCD1LG2 PLCB4 POLD1 PPARG PPP2R2A RPS6KA3 SDHA SDHC TCF7L2 VEGFA WRN WWTR1 YAP1*.

Starting from 100 to 250 ng of genomic DNA, enzymatic (KAPA HyperPlus library preparation kit, Roche, Pleasanton, CA) or mechanical (ME220 Focused-ultrasonicator, Covaris, Woburn, MA) fragmentation was performed, followed by library preparation (KAPA HyperPlus library preparation kit). DNA libraries were enriched by hybridization capture using xGen Lockdown

Probes (Integrated DNA Technologies, Coralville, IA), and sequenced on a MiSeq or NextSeq 550 system (Illumina, San Diego, CA).

**Mutation calling and mutational signature analysis**

Single-nucleotide variants (SNVs) were called using Mutect2, and germline single-nucleotide polymorphisms (SNPs) with HaplotypeCaller. The somatic mutations were selected by filtering out HaplotypeCaller mutations and common variants with MAF > 0.1% (ExAC, gnomAD, and 1000 genomes) annotated using Annovar, and low variant-allele frequency variants or those with low coverage were excluded. These variants were only used in signature analysis. The specific gene-level variants reported in the manuscript were obtained after evaluation by the pathology department.

The 96-dimensional mutational spectra were constructed by counting mutations based on 3-nucleotide context and substitution type, and Sig3 was detected using pre-trained (data = 'msk') classifiers in SigMA (24) with corresponding tumor type options ('ovary', 'panc_ad', 'prost', 'head', 'colorectal', 'biliary'. The CR patient with high-grade serous ovarian carcinoma had a SigMA score of 0.92, passing the strict 5% false-positive rate threshold.

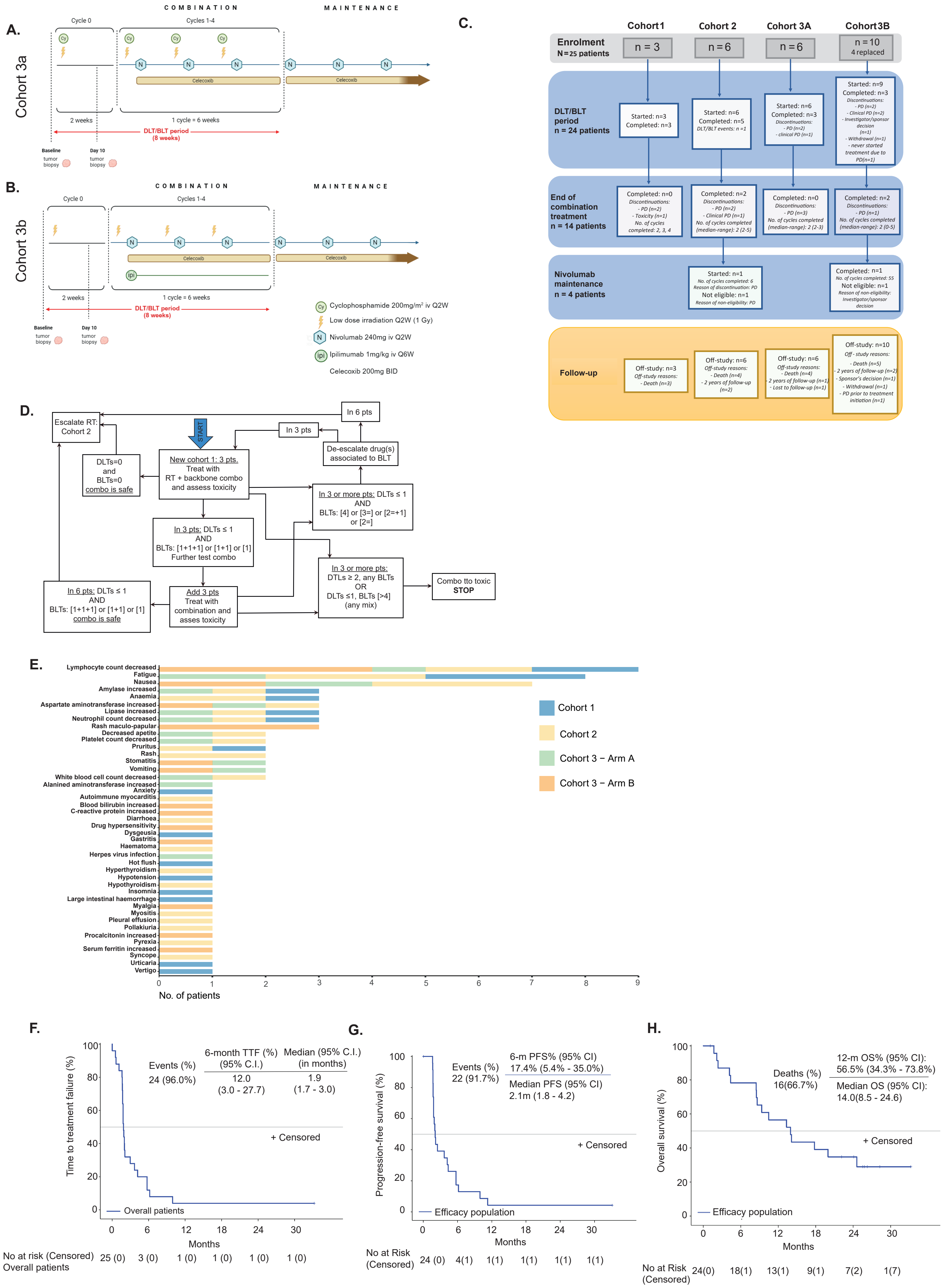

### Supplement 1

**Supp. 1:**

Therapeutic schema of the phase I clinical trial RACIN for cohort 3, arm **(A)** A and **(B)** B. **(C)** Consort flowchart of patients included in the clinical trial RACIN. **(D)** Adapted 3+3 algorithm for multidrug DLT/BLT assessment.in cohort 1. **(E)** Stacked barchart depicting frequency of treatment-related toxicities in the DLT/BLT period, by cohort/arm. **(F)** Kaplan-Meier estimate of time to treatment failure (TTF). Treatment failure includes discontinuation due to progression, toxicity, death, investigator/sponsor decision, withdrawal or lost to follow-up (whichever comes first). Kaplan-Meier estimate of **(G)** Progression Free Survival and **(H)** Overall Survival.

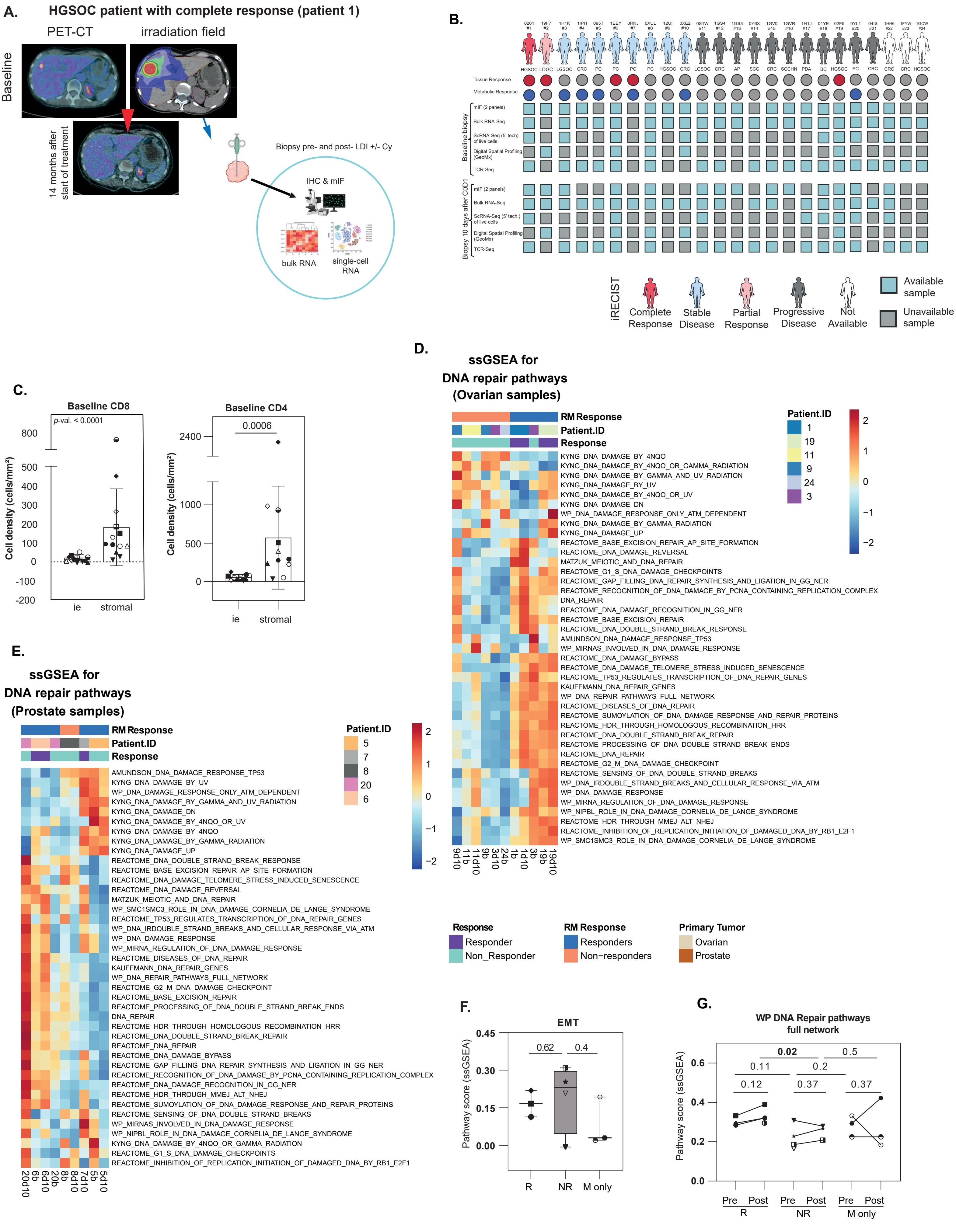

Supplement 2

**<u>Supp 2:</u> (A)** Workflow of translational analysis performed, exemplified with PET/CT scan and irradiation field of a HGSOC patient achieving CR. Patients underwent paired biopsies of irradiated metastases at baseline and 10 days (+/- 3 days) following LDRT +/- Cy, but before the initiation of ICB. Immunohistochemistry (IHC), multiplex immunofluorescence (mIF), bulk RNA, and single-cell RNA were used to provide a comprehensive analysis of the TME and of LDRT-induced changes in immune-excluded tumors. Post-treatment changes in the TME assessed solely the activity of LDRT +/- Cy, as ICB was administered after the day 10 biopsy. **(B)** Sample availability per patient and per timepoint. Available material is depicted in light blue, while unavailable material appears in grey. **(C)** Left: Box plot displaying cell density (cells/mm$^2$) of intraepithelial and stromal CD8$^+$ T cells of all patients by mIF at baseline (Mann-Whitney, two-tailed). Right: Box plot showing cell density (cells/mm$^2$) of intraepithelial and stromal CD4$^+$ T cells of all patients by mIF at baseline (Mann-Whitney, two-tailed). Heatmaps showing select DNA repair pathways in ovarian **(D)** and prostate **(E)** samples. The heatmaps display ssGSEA scores for reactome DNA repair pathways from bulk RNA sequencing. **(F)** Box plot displaying ssGSEA pathway scores from bulk RNA Seq at baseline for EMT. **(G)** Line graphs showing ssGSEA pathway scores from bulk RNA Seq at baseline and at day 10 for different DNA repair pathway signatures (paired: Wilcoxon test; unpaired: Mann-Whitney test; one tailed).

LDGC: low differentiated gallbladder carcinoma. HGSOC: high grade serous ovarian carcinoma. PC: prostate adenocarcinoma. CRC: Colorectal adenocarcinoma. LGSOC: low grade serous ovarian carcinoma. SCC: Squamous cervical carcinoma. SCCHN: squamous cell carcinoma of the head and neck. PDA: pancreatic ductal adenocarcinoma. BC: ductal breast carcinoma. AP: appendix adenocarcinoma. Sample IDs: b: baseline sample (pre-treatment); d10: day 10 sample (post-treatment).

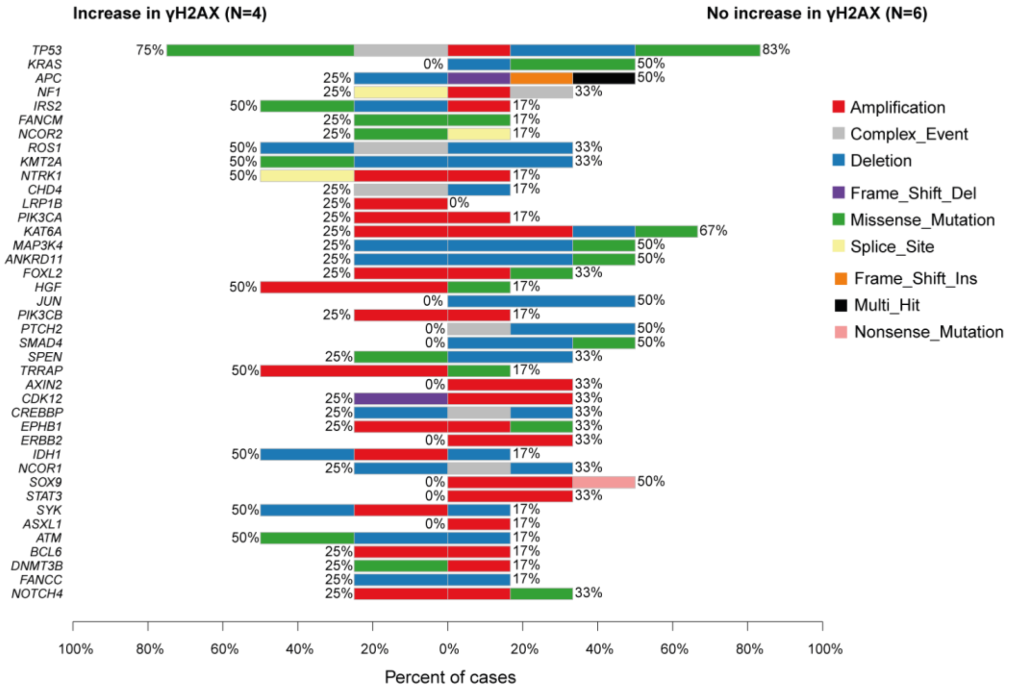

#### Supplement 3

**<u>Supp. 3</u>**: Bar graph with mutation frequencies obtained from an 400-gene high-throughput sequencing (HTS) panel, categorized by increase in γH2AX (left) or no increase in γH2AX (right) following LDRT +/- Cy.

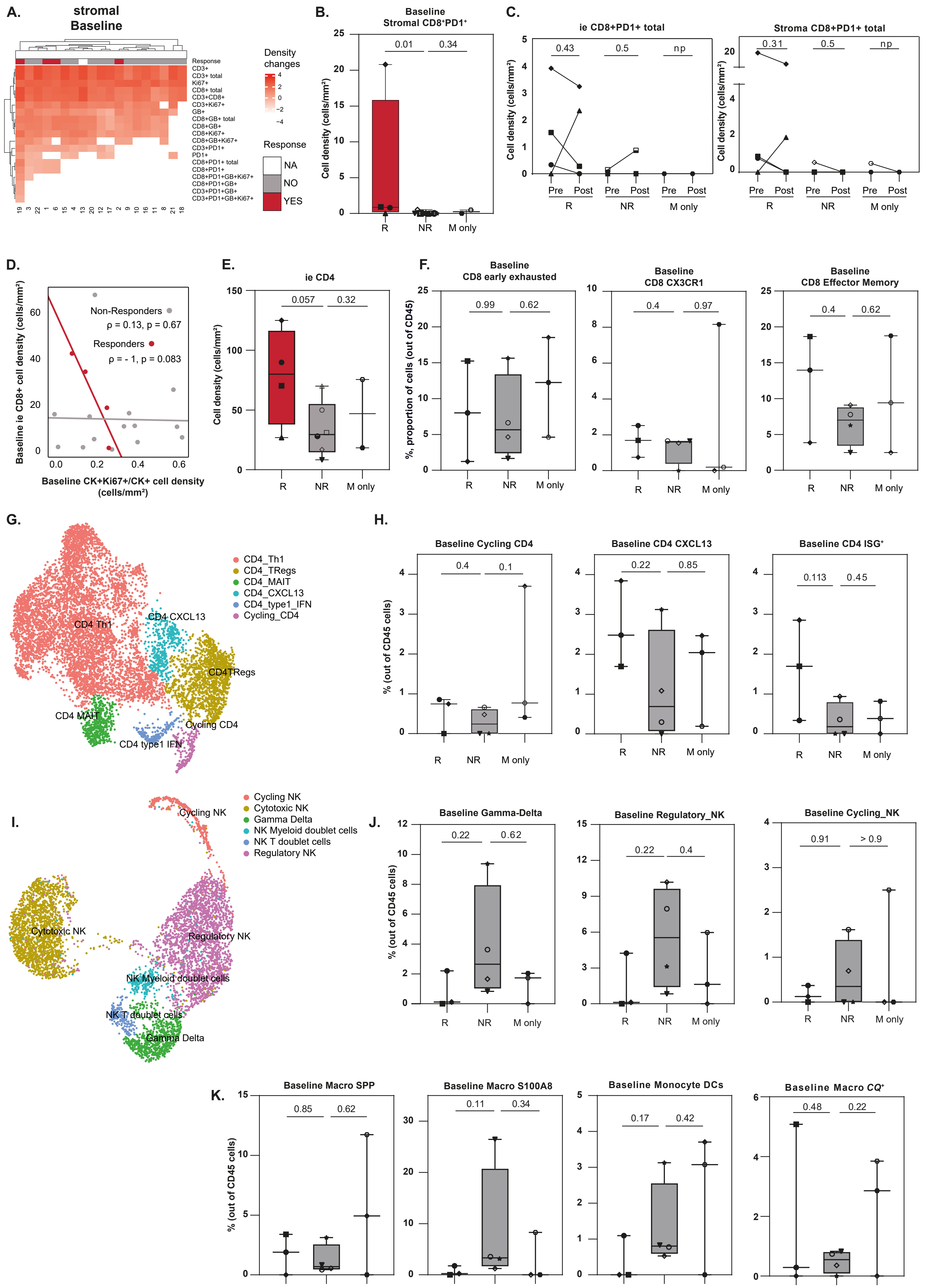

### Supplement 4

**Supp. 4:**

**(A)** Heatmap of stromal cell densities quantified by mIF of different T cell phenotypes in baseline tumor biopsies. **(B)** Boxplot displaying cell density (cells/$mm^2$) of stromal CD8+PD1+ cells at baseline by mIF (Mann-Whitney, two-tailed). **(C)** Line graph displaying cell densities (cells/mm2) of intraepithelial and stromal $CD8^+PD1^+$ T cells at baseline and at day 10 by mIF (pre vs post: Wilcoxon test, one tailed). **(D)** Scatter plot displaying Spearman rank correlation of intraepithelialCD8+ T cell density (cells/$mm^2$) and proportion of proliferating tumor cells (defined by fraction of proliferating tumors cells, $CK^+Ki67^+/CK^+$) at baseline, done by mIF (Responders: $\rho = -1$, p value 0.083). Low tumor cell proliferation correlates with high CD8+ T cell infiltration in tumor nests in R patients, suggesting that in this subgroup of patients, the lower the tumor aggressiveness, the higher the $CD8^+$ T cell infiltration in the tumor. **(E)** Boxplot showing cell density (cells/$mm^2$) of intraepithelialCD4 at baseline by mIF (Mann-Whitney, two-tailed). **(F)** Boxplot displaying proportion of $CD8^+$ early exhausted, CX3CR1 and effector memory T cells in the $CD45^+$ cells at baseline (Mann-Whitney, two-tailed). **(G)** UMAP projection displaying sub-clustering of $CD4^+$ T cells from viable-sorted data of ten baseline and eight post-treatment tumors. **(H)** Boxplot displaying proportion of cycling CD4, CD4 CXCL13 and CD4 $ISG^+$ T cells in the $CD45^+$ cells at baseline (Mann-Whitney, two-tailed). **(I)** UMAP projection displaying sub-clustering of innate T cells from viable-sorted data of ten baseline and eight post-treatment tumors. **(J)** Boxplots displaying proportion of gamma-delta, regulatory and cycling NK cells in the $CD45^+$ cells at baseline (Mann-Whitney, one-tailed). **(K)** Boxplots displaying proportion of different cells from the myeloid compartment in the $CD45^+$ cells at baseline (Mann-Whitney, one-tailed).

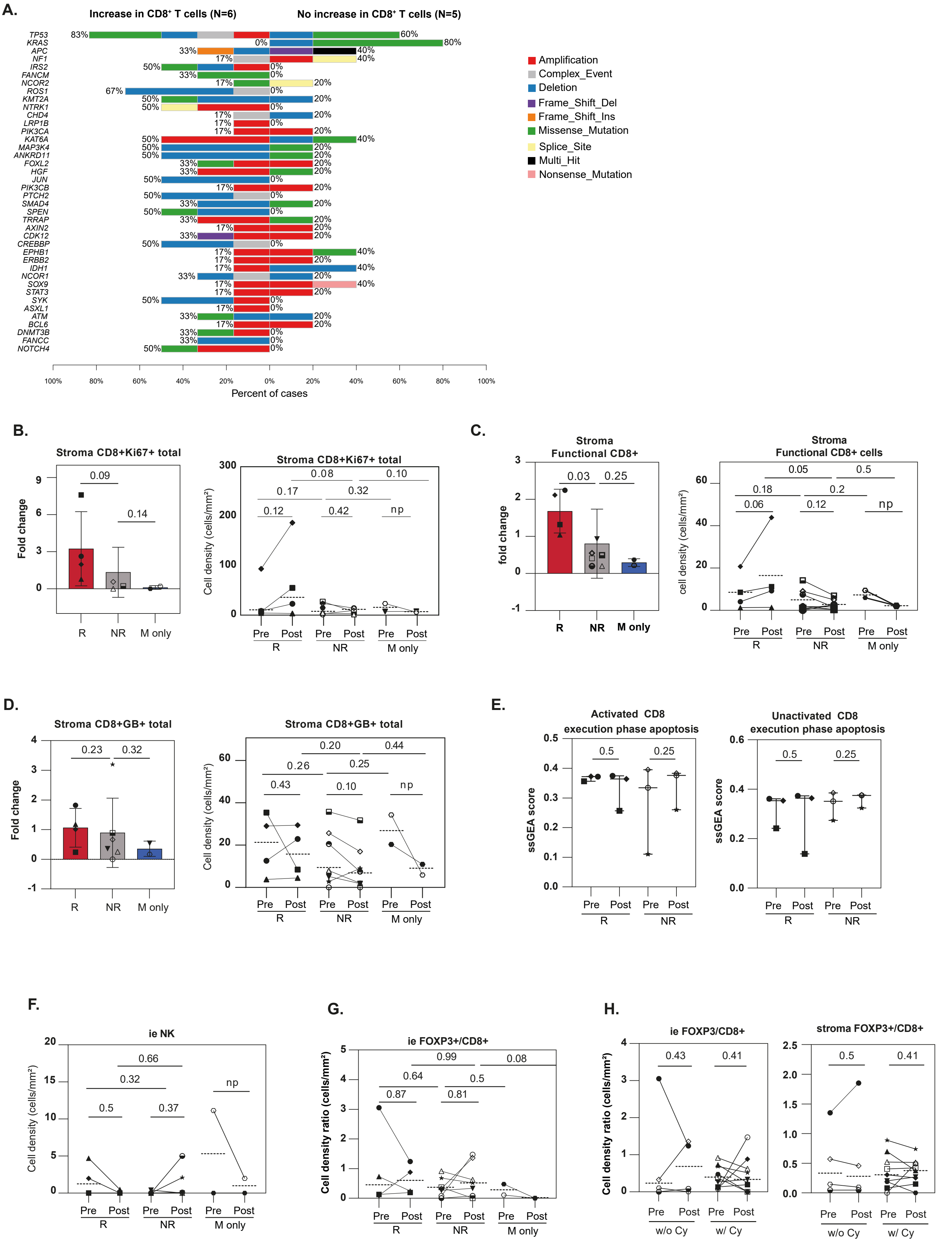

**Supplement 5**

**Supp. 5:**

**(A)** Bar graph with mutation frequencies obtained from an 400-gene high-throughput sequencing (HTS) panel, categorized by increase in $CD8^+$ T cells (left) or no increase in $CD8^+$ T cells (right) following LDRT +/- Cy. **(B)** Left: Box plot showing fold change of proliferating stromal CD8+Ki67+ T cells after LDRT +/- Cy by mIF (Mann-Whitney, one-tailed). Right: Line graph showing cell densities (cells/mm2) of proliferating stomal $CD8^+Ki67^+$ T cells at baseline and at day 10 by mIF (pre vs post: Wilcoxon, one-tailed; rest: Mann-Whitney, one-tailed) **(C)** Left: Box plot showing fold change of functional stromal $CD8^+$ T cells after LDRT +/- Cy by mIF (Mann-Whitney, one-tailed). Right: Line graph showing cell densities (cells/mm2) of functional stomal $CD8^+$ T cells at baseline and at day 10 by mIF (pre vs post: Wilcoxon, one-tailed; rest: Mann-Whitney, one-tailed). **(D)** Left: Box plot showing fold change of cytotoxic stromal $CD8^+GB^+$ T cells after LDRT +/- Cy by mIF (Mann-Whitney, one-tailed). Right: Line graph showing cell densities (cells/mm2) of cytotoxic stromal $CD8^+GB^+$ T cells at baseline and at day 10 by mIF (pre vs post: Wilcoxon, one-tailed; rest: Mann-Whitney, one-tailed). **(E)** Box plot displaying pathway score (ssGSEA) from scRNA seq of the pathway execution phase apoptosis in activated and unactivated $CD8^+$ T cells (Wilcoxon, one-tailed) **(F)** Line graph displaying cell density (cells/mm2) of intraepithelial NK cells at baseline and at day 10 by mIF (NB: many patients did not display NK cells per mIF) (pre vs post: Wilcoxon, one-tailed; rest: Mann-Whitney, one-tailed) **(G)** Line graph displaying cell density ratio of intraepithelial $FOXP3^+/CD8^+$ cells at baseline and at day 10 by mIF (pre vs post: Wilcoxon, one-tailed; rest: Mann-Whitney, one-tailed). **(H)** Line graphs displaying cell density ratio of intraepithelial and stromal $FOXP3^+/CD8^+$ cells at baseline and at day 10, categorized by patients not having received Cyclophosphamide (w/o Cy) vs having received Cyclophosphamide (w/Cy) (Wilcoxon test, one-tailed).

| | Cohort 1 | Cohort 2 | Cohort 3 - Arm A | Cohort 3 - Arm B |
|---|---|---|---|---|
| **Enrolled patients** | **3** | **6** | **6** | **10** |
| **Patients started treatment** | **3** | **6** | **6** | **9**[1] |
| **Replaced patients** | **-** | **-** | **-** | 4[2] |
| **Low dose ionizing radiation (Cohort 1: 0.5Gy; Cohorts 2-3(A&B): 1Gy) - Q2W** | | | | |
| No. of patients started | 3 | 6 | 6 | 9 |
| No. of patients completed | - | 2[3] | - | 2 |
| No. of doses received (median - range) | *No. of doses:* 6, 7, 10 | 6 (4 – 13) | 4 (4 – 7) | 4 (1 – 13) |
| **Cyclophosphamide (200 mg/m2) IV - Q2W** | | | | |
| No. of patients started | 3 | 6 | 6 | *NA* |
| No. of patients completed | - | 2 | - | *NA* |
| No. of doses received (median - range) | *No. of doses:* 6, 7, 10 | 5 (2 – 13) | 4 (2 – 7) | *NA* |
| **Nivolumab in combination treatment (240 mg) IV - Q2W** | | | | |
| No. of patients started | 3 | 6 | 6 | 7 |
| No. of patients completed | - | 2[4] | - | 2[4] |
| No. of doses received (median - range) | *No. of doses:* 5, 6, 9 | 5 (3 – 12) | 3 (3 – 6) | 3 (1 – 12) |
| **Ipilimumab (1 mg/Kg) IV - Q6W** | | | | |
| No. of patients started | 3 | 6 | *NA* | 7 |
| No. of patients completed | - | - | *NA* | 3 |
| No. of doses received (median - range) | *No. of doses:* 2 for all pts | 2 (1 – 2) | *NA* | 1 (1 – 4) |
| **Aspirin (300 mg/day) P.O.** | | | | |
| No. of patients started | 3 | 4 | *NA* | *NA* |
| No. of patients completed | - | - | *NA* | *NA* |
| **Celecoxib (200 mg BID) P.O.** | | | | |
| No. of patients started | *NA* | *NA* | 5 | 7 |
| No. of patients completed | *NA* | *NA* | - | 2 |
| **End of DLT/BLT period (C0D1 to C2D1(pre-dose))** | | | | |
| No. of patients evaluable for MTD | 3 | 6 | 6 | 6 |
| No. of patients completed without any limiting toxicity (DLT/BLT event) | 3 | 5[5] | 3[6] | 3 |
| No. of patients who had a DLT or a BLT event | - | 1[7] | - | - |
| No. of patients who failed (after C1D30) → no further treatment administration | - | - | 3 | 3 |
| No. of patients who failed before C1D30 → replaced patients | - | - | - | 4 |
| *Reasons of combination treatment failure:* | | | | |
| Radiological progression | - | - | 2 | 3 |
| Clinical progression | - | - | 1 | 2 |
| Toxicity (BLT event) | - | 17 | - | - |
| Investigator/sponsor decision | - | - | - | 1 |
| Withdrawal by subject | - | - | - | 1 |
| **End of combination treatment (C0D1 to C4D30)** | | | | |
| No. of patients completed | - | 2 | - | 2 |

| | | | | |
|---|---|---|---|---|
| No. of cycles completed (median - range) | *No. of cycles:* 2, 3, 4 | 2 (2 – 5) | 2 (2 – 3) | 2 (0 – 5) |
| *Reasons of combination treatment failure after completion of DLT/BLT period:* | | | | |
| Radiological progression | 2 | 2 | 3 | 1 |
| Clinical progression | - | 1 | - | - |
| Toxicity | 1[8] | - | - | - |
| **Eligibility for nivolumab maintenance** | | | | |
| No. of eligible patients | - | 1 | - | 1 |
| No. of patients who completed the combination treatment but considered ineligible for nivolumab maintenance | - | 1 | - | 1 |
| *Reasons of non-eligibility:* | | | | |
| *Radiological progression* | - | 1 | - | - |
| Investigator/sponsor decision | - | - | - | 1 |
| **Nivolumab maintenance (240 mg) IV - Q2W** | | | | |
| No. of patients started | - | 1 | - | 1 |
| No. of doses/cycles received | - | 6 | - | 23 |
| *Reasons of nivolumab maintenance failure:* | | | | |
| Disease progression | - | 1 | - | - |
| **Follow-up** | | | | |
| No. of patients still on treatment | - | - | - | 1 |
| No. of patients on survival follow-up | - | - | 2 | 4 |
| No. of patients off-study | 3 | 6 | 4 | 5 |
| *Off-study reasons:* | | | | |
| Death | 3 | 4 | 4 | 3 *(2 replaced)* |
| *Two years of follow-up (post-treatment start)* | - | 2 | - | - |
| Withdrawal by subject | - | - | - | 1 *(replaced)* |
| *Progression prior to treatment initiation* | - | - | - | 1 *(replaced)* |

*(1) Patient had PD one week post-randomisation, and went off-study without having received any treatment.*
*(2) Four patients were replaced due to treatment failure prior to completion of the DLT/BLT evaluation period for other reason than a DLT/BLT:*
*- One patient had clinical PD 2.5 weeks post-randomisation and went off-treatment.*
*- One patient withdrew consent five weeks post-randomisation.*
*- One patient had PD one week post-randomisation, and went off-study without receiving any treatment.*
*- One patient discontinued treatment one week post-randomisation due to investigator decision. Note that replaced patients are not included in the MTD population.*
*(3) For one patient, one radiotherapy dose skipped.*
*(4) For one patient, one nivolumab dose skipped.*
*(5) One patient had PD in the DLT/BLT period but continued treatment and completed the DLT/BLT period.*
*(6) Two patients had PD in the DLT/BLT period but continued treatment and completed the DLT/BLT period.*
*(7) Autoimmune myocarditis of G4 (also a BLT event and a SAE), probably related to nivolumab and ipilimumab, and possibly related to cyclophosphamide.*
*(8) Immune related colitis of G3 (also a SAE), probably related to nivolumab and ipilimumab.*

**Supp. Table 1**: Information on treatment phases and follow-up, by cohort/arm

| DCR at 3 months (95% binomial C.I.) | DCR at 6 months (95% binomial C.I.) | DCR at 12 months (95% binomial C.I.) |
|---|---|---|
| **Cohort 1 (n= 3 patients)** | | |
| 66.7% (9.4% - 99.2%) | 0.0% (0.0% - 70.8%)[b] | 0.0% (0.0% - 70.8%)[b] |
| **Cohort 2 (n= 6 patients)** | | |
| 33.3% (4.3% - 77.7%) | 16.7% (0.4% - 64.1%) | 0.0% (0.0% - 45.9%)[b] |
| **Cohort 3 - Arm A (n= 6 patients)** | | |
| 0.0% (0.0% - 45.9%)[b] | 0.0% (0.0% - 45.9%)[b] | 0.0% (0.0% - 45.9%)[b] |
| **Cohort 3 - Arm B (n= 9 patients)** | | |
| 22.2% (2.8% - 60.0%) | 11.1% (0.3% - 48.2%) | 11.1% (0.3% - 48.2%) |

*(a) Complete, partial response or stable disease.*

*(b) 95% Clopper Pearson exact C.I.*

**Supp. Table 2:** Disease control[a] rate (DCR) by RECIST 1.1 (and by PCWG3 for prostate cancer), by arm/cohort for the efficacy population (n = 24)

| No. of patients | No. of PFS events (%) | Median (95% C.I.) PFS (in months) |
|---|---|---|
| **Cohort 1** | | |
| 3 | 3 (100.0) | 5.7 (1.9 - 11.3) |
| **Cohort 2** | | |
| 6 | 6 (100.0) | 4.0 (1.7 – 10.0) |
| **Cohort 3A** | | |
| 6 | 6 (100.0) | 1.8 (1.6 – 2.1) |
| **Cohort 3B** | | |
| 9 | 7 (77.8) | 2.1 (1.7 - 6.2) |

**Supp. Table 3:** Progression free survival (PFS) by RECIST 1.1 (and by PCWG3 for prostate cancer), by arm/cohort for the efficacy population (n = 24)

| No. of patients | No. of deaths (%) | Median (95% C.I.) OS (in months) |
|---|---|---|
| **Cohort 1** | | |
| 3 | 3 (100.0) | 14.1 (8.5 – 24.6) |
| **Cohort 2** | | |
| 6 | 4 (66.7) | 13.7 (4.4 – NE) |
| **Cohort 3 - Arm A** | | |
| 6 | 4 (66.7) | 14.1 (4.2 – NE) |
| **Cohort 3 - Arm B** | | |
| 9 | 3 (33.3) | NR (1.7 – NE) |

*NR: Not reached; NE: Not evaluable*

**Supp. Table 4:** Overall survival (OS), by arm/cohort for the efficacy population (n = 24)

The following DLTs/BLTs were to be attributed to one or more components of the treatment:

**A. Toxicity related to the combination of study treatments: BLTs group A**

- Grade 4-5 allergic reactions related to the infusion of any of the products in the study, extending for more than 2 hours after infusion, and not reversible to a grade 2 or less within 24 hours of infusion administration with standard therapy.
- Grade ≥4 autoimmune reactions.
- Grade ≥4 major organ toxicity (cardiac, pulmonary, hepatic, renal, neurological) not pre-existing and not related to the underlying malignancy, not resolving to grade ≤ 2 within 14 days, and occurring within the DLT/BLT period after infusion of any of the products in the study.
- Any AEs related to the drug combination (or that cannot be clearly assigned to any of the other components of the backbone treatment) leading to permanent discontinuation of treatment, as per protocol criteria.

**B. Toxicity related to cyclophosphamide: BLTs group B**

- Grade ≥4 neutropenia (ANC < 500/µL)
- Grade ≥3 febrile neutropenia lasting ≥ 10 days
- Grade ≥4 thrombocytopenia lasting > 5 days
- Any AEs related to cyclophosphamide leading to permanent discontinuation of treatment, as per protocol criteria.

**C. Toxicity related to aspirin: BLTs group C**

- Grade ≥2 hemorrhagic episodes
- Grade ≥2 gastritis or peptic ulcer
- Any AEs related to aspirin leading to permanent discontinuation of treatment, as per protocol criteria.

**D. Toxicity related to celecoxib: BLT group C (For Cohort 3, 4 and Phase Ib)**

- Grade ≥2 hemorrhagic episodes

- Grade ≥2 gastritis or peptic ulcer

**E. Toxicity related to radiation2 (group D): DLTs**

- Grade >3 local reaction that appears inside the irradiated volume (i.e. esophagitis if the esophagus was in the treatment field)

**Supp. Table 5: DLT/BLT definitions**
